# Large scale genome-wide association analyses identify novel genetic loci and mechanisms in hypertrophic cardiomyopathy

**DOI:** 10.1101/2023.01.28.23285147

**Authors:** Rafik Tadros, Sean L Zheng, Christopher Grace, Paloma Jordà, Catherine Francis, Sean J Jurgens, Kate L Thomson, Andrew R Harper, Elizabeth Ormondroyd, Dominique M West, Xiao Xu, Pantazis I Theotokis, Rachel J Buchan, Kathryn A McGurk, Francesco Mazzarotto, Beatrice Boschi, Elisabetta Pelo, Michael Lee, Michela Noseda, Amanda Varnava, Alexa MC Vermeer, Roddy Walsh, Ahmad S Amin, Marjon A van Slegtenhorst, Nicole Roslin, Lisa J Strug, Erika Salvi, Chiara Lanzani, Antonio de Marvao, Hypergenes InterOmics Collaborators, Jason D Roberts, Maxime Tremblay-Gravel, Genevieve Giraldeau, Julia Cadrin-Tourigny, Philippe L L’Allier, Patrick Garceau, Mario Talajic, Yigal M Pinto, Harry Rakowski, Antonis Pantazis, John Baksi, Brian P Halliday, Sanjay K Prasad, Paul JR Barton, Declan P O’Regan, Stuart A Cook, Rudolf A de Boer, Imke Christiaans, Michelle Michels, Christopher M Kramer, Carolyn Y Ho, Stefan Neubauer, HCMR Investigators, Paul M Matthews, Arthur A Wilde, Jean-Claude Tardif, Iacopo Olivotto, Arnon Adler, Anuj Goel, James S Ware, Connie R Bezzina, Hugh Watkins

**Affiliations:** Cardiovascular Genetics Centre, Montreal Heart Institute, Montreal, QC, Canada; Faculty of Medicine, Université de Montréal, Montreal, QC, Canada; Department of Experimental Cardiology, Amsterdam Cardiovascular Sciences, University of Amsterdam, Amsterdam UMC, Amsterdam, the Netherlands; National Heart & Lung Institute, Imperial College London, London, UK; MRC London Institute of Medical Sciences, Imperial College London, London, UK; Royal Brompton & Harefield Hospitals, Guy’s and St. Thomas’ NHS Foundation Trust, London, UK; Radcliffe Department of Medicine, University of Oxford, Division of Cardiovascular Medicine, John Radcliffe Hospital, Oxford, UK; Wellcome Centre for Human Genetics, University of Oxford, Oxford, UK; Cardiovascular Disease Initiative, Broad Institute of MIT and Harvard, Cambridge, MA, USA; Oxford Genetics Laboratories, Churchill Hospital, Oxford, UK; Department of Molecular and Translational Medicine, University of Brescia, Brescia, Italy; Genetics Unit, Careggi University Hospital, Florence, Italy; Imperial College Healthcare NHS Trust, Imperial College London, London, UK; Department of Clinical Genetics, University of Amsterdam, Amsterdam UMC, Amsterdam, the Netherlands; European Reference Network for Rare and Low Prevalence Complex Diseases of the Heart, (ERN GUARD-HEART; https://guardheart.ern-net.eu); Department of Clinical Cardiology, Amsterdam Cardiovascular Sciences, University of Amsterdam, Amsterdam UMC, Amsterdam, the Netherlands; Department of Clinical Genetics, Erasmus Medical Center, University Medical Center Rotterdam, Rotterdam, the Netherlands; The Centre for Applied Genomics, Genetics and Genome Biology, The Hospital for Sick Children, Toronto, ON, Canada; Departments of Statistical Sciences and Computer Science, Data Sciences Institute, University of Toronto, Toronto, ON, Canada; The Centre for Applied Genomics, The Hospital for Sick Children, Toronto, ON, Canada; Ontario Regional Centre, Canadian Statistical Sciences Institute, University of Toronto, Toronto, ON, Canada; Neuroalgology Unit, Fondazione IRCCS Istituto Neurologico Carlo Besta, Milan, Italy; Genomics of Renal Diseases and Hypertension Unit, Nephrology Operative Unit, IRCCS San Raffaele Hospital, Milan, Italy; Chair of Nephrology, Vita-Salute San Raffaele University, Milan, Italy; Section of Cardiac Electrophysiology, Division of Cardiology, Department of Medicine, Western University, London, ON, Canada; Peter Munk Cardiac Centre, Toronto, ON, Canada; National Heart Centre Singapore, Singapore; Duke-National University of Singapore Medical School, Singapore; Department of Cardiology, Thoraxcenter, Erasmus University Medical Center, Rotterdam, the Netherlands; Department of Genetics, University of Groningen, University Medical Center Groningen, Groningen, the Netherlands; Department of Medicine, Cardiovascular Division, University of Virginia Health, Charlottesville, VA, USA; Cardiovascular Division, Brigham and Women’s Hospital, Boston, MA, USA; Division of Cardiovascular Medicine, Radcliffe Department of Medicine, NIHR Oxford Health Biomedical Research Centre, University of Oxford, Oxford, UK; Department of Brain Sciences and UK Dementia Research Institute, Imperial College London, London, UK; ECGen, Cardiogenetics Focus Group of EHRA, France; Department of Experimental and Clinical Medicine, Meyer Children Hospital, University of Florence, Florence, Italy; Division of Cardiology, Peter Munk Cardiac Centre, University Health Network, Toronto, ON, Canada; Department of Medicine, University of Toronto, Toronto, ON, Canada; Program in Medical & Population Genetics, Broad Institute of MIT and Harvard, Cambridge, MA, USA

**Author notes:** These authors contributed equally to this work (Rafik Tadros, Sean L Zheng, Anuj Goel, James S Ware, Connie R Bezzina, Hugh Watkins). <u>Hypergenes InterOmics Collaborators</u>: Daniele Cusi, Paolo Manunta, Lorena Citterio, Nicola Glorioso. <u>HCMR Investigators</u>: Theodore Abraham, Lisa Anderson, Evan Appelbaum, Camillo Autore, Colin Berry, Elena Biagini, William Bradlow, Chiara Bucciarelli-Ducci, Amedeo Chiribiri, Lubna Choudhury, Andrew Crean, Dana Dawson, Milind Desai, Patrice Desvigne-Nickens, Eleanor Elstein, Andrew Flett, Matthias Friedrich, Nancy Geller, Stephen Heitner, Adam Helms, Daniel Jacoby, Dong-Yun Kim, Han Kim, Bette Kim, Eric Larose, Masliza Mahmod, Heiko Mahrholdt, Martin Maron, Gerry McCann, Saidi Mohiddin, Francois-Pierre Mongeon, Sherif Nagueh, David Newby, Anjali Owens, Sven Plein, Ornella Rimoldi, Michael Salerno, Jeanette Schulz-Menger, Mark Sherrid, Albert van Rossum, Jonathan Weinsaft, James White, Eric Williamson.

## Abstract

Hypertrophic cardiomyopathy (HCM) is an important cause of morbidity and mortality with both monogenic and polygenic components. We here report results from the largest HCM genome-wide association study (GWAS) and multi-trait analysis (MTAG) including 5,900 HCM cases, 68,359 controls, and 36,083 UK Biobank (UKB) participants with cardiac magnetic resonance (CMR) imaging. We identified a total of 70 loci (50 novel) associated with HCM, and 62 loci (32 novel) associated with relevant left ventricular (LV) structural or functional traits. Amongst the common variant HCM loci, we identify a novel HCM disease gene, *SVIL*, which encodes the actin-binding protein supervillin, showing that rare truncating *SVIL* variants cause HCM. Mendelian randomization analyses support a causal role of increased LV contractility in both obstructive and non-obstructive forms of HCM, suggesting common disease mechanisms and anticipating shared response to therapy. Taken together, the findings significantly increase our understanding of the genetic basis and molecular mechanisms of HCM, with potential implications for disease management.

---

HCM is a disease of cardiac muscle characterized by thickening of the LV wall with an increased risk of arrhythmia, heart failure, stroke and sudden death. Previously viewed as a Mendelian disease with rare pathogenic variants in cardiac sarcomere genes identified in ∼35% of cases (HCM_SARC+_), HCM is now known to have complex and diverse genetic architectures.^1^ Prior studies have established that common genetic variants underlie a large portion of disease heritability in HCM not caused by rare pathogenic variants (HCM_SARC-_) and partly explain the variable expressivity in HCM patients carrying pathogenic variants (HCM_SARC+_), but such studies had limited power to identify a large number of significant loci.^2,3^

We report a new meta-analysis of 7 case-control HCM GWAS datasets, including 3 not previously published, comprising a total of 5,900 HCM cases, 68,359 controls and 9,492,702 variants with a minor allele frequency (MAF)>1% (**Supplementary Table 1**; Study flowchart in **Figure 1**). Using the conventional genome-wide significance threshold (P< 5×10^−8^), 34 loci were significantly associated with HCM, of which 15 were novel (**Table 1**). We then performed 2 stratified analyses in HCM_SARC+_ (1,776 cases) and HCM_SARC-_ (3,860 cases), and identified an additional 1 locus and 4 loci, respectively (**Table 1**; **Supplementary Table 2**; **Supplementary Figure 1**). Using conditional analysis^4^, we identified additional suggestive and independent associations with HCM, HCM_SARC+_, and HCM_SARC-_ with a false discovery rate (FDR) <1% (**Supplementary Table 3**). A locus on chromosome 11 which includes *MYBPC3*, a well-established disease gene, is associated with HCM and HCM_SARC+_, but not HCM_SARC-_, implying that this association is tagging known founder pathogenic variants in *MYBPC3*.^2,3^ We estimated the heritability of HCM attributable to common genetic variation (*h*^2^_SNP_) in the all-comer analysis to be 0.17±0.02 using LD score regression (LDSC)^5^, and, as expected, found higher estimates (0.25±0.02) using genome-based restricted maximum likelihood (GREML)^6^, with higher *h*^2^_SNP_ in HCM_SARC-_ (0.29±0.02) compared to HCM_SARC+_ (0.16±0.04) (**Supplementary Table 4**).

**Figure 1:**
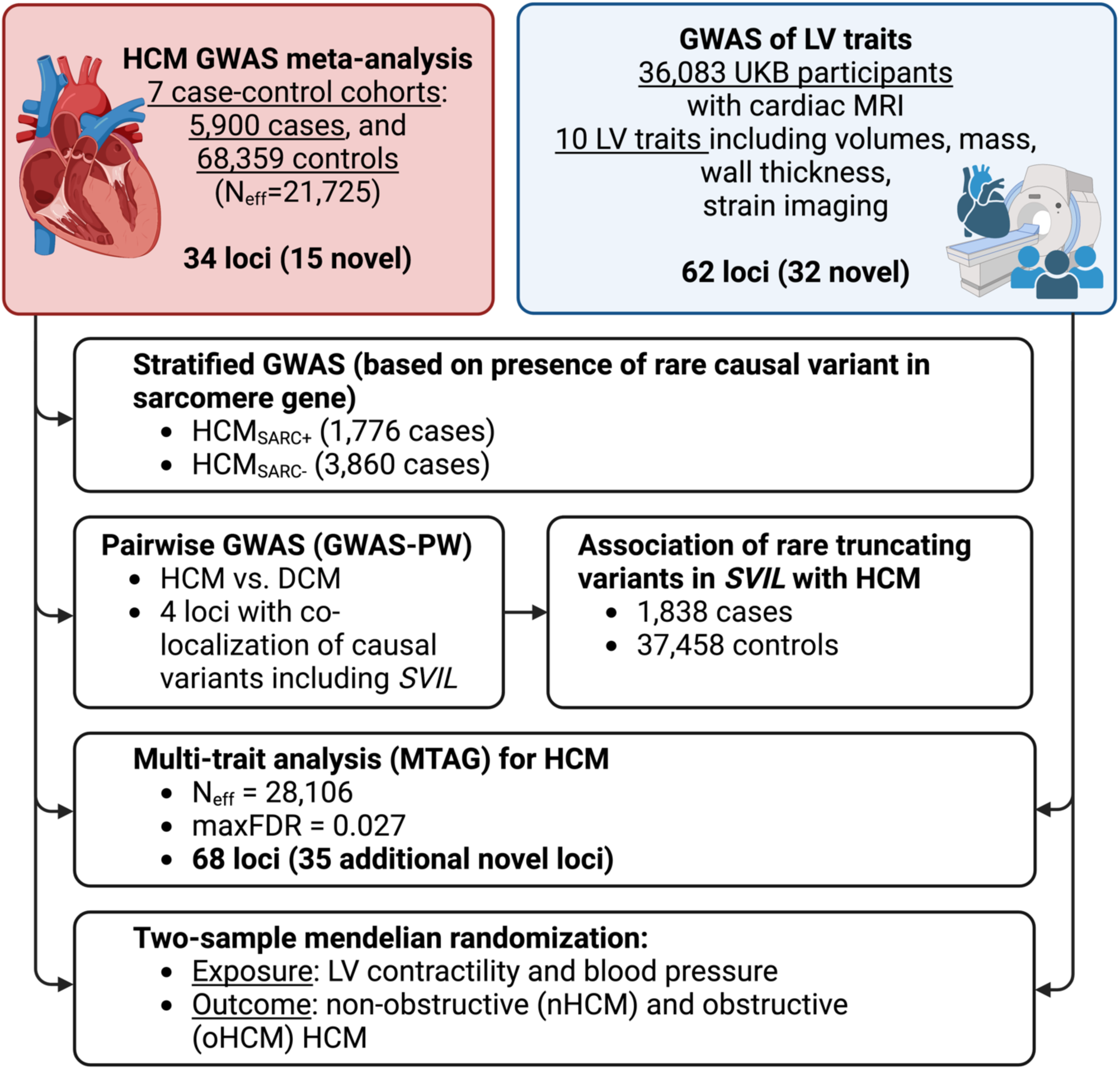
Study Flowchart. Abbreviations: DCM, dilated cardiomyopathy; HCM, hypertrophic cardiomyopathy; LV, left ventricular; maxFDR, upper bound of the estimated false discovery rate computed using MTAG; MRI, magnetic resonance imaging; N_eff_, effective sample size (see methods); UKB, UK Biobank.

**Table 1:**
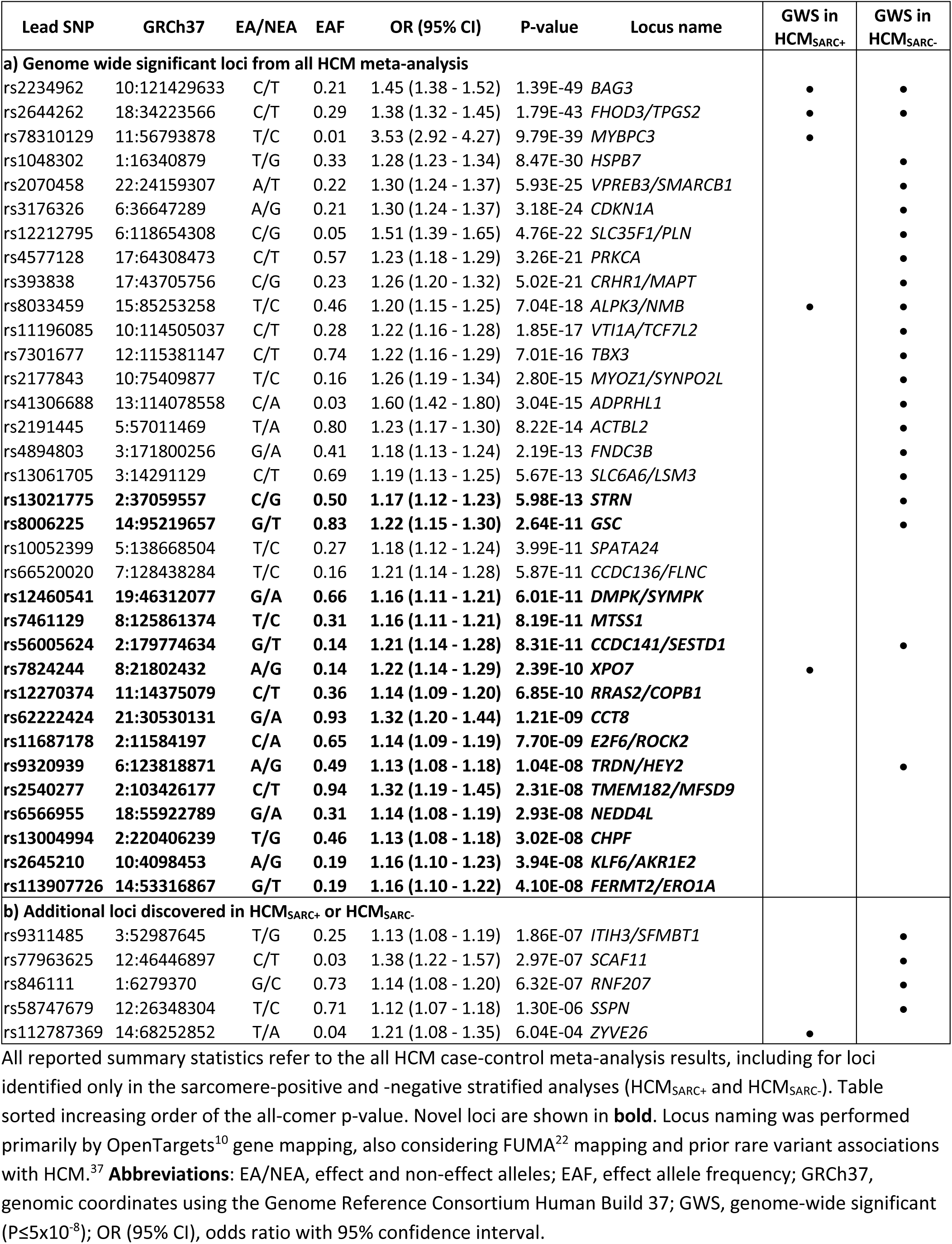
Lead variants from the HCM meta-analysis.

Rare variants in sarcomere genes that cause HCM and dilated cardiomyopathy (DCM) are known to have opposing effects on contractility^7^ and we previously demonstrated that HCM and DCM GWAS loci similarly overlap, with opposite direction of effect.^3^ We leveraged such opposing genomic effects in HCM and DCM to identify additional loci involved in HCM. Bayesian pairwise analysis (GWAS-PW^8^) including the present HCM GWAS meta-analysis and a published DCM GWAS^9^ identified four genomic regions where the same variant was deemed causal for both diseases with a posterior probability >0.9 (**Supplementary Table 5**). In all 4 genomic regions, opposing directional effects were observed in HCM and DCM. The top mapped genes at these loci using OpenTargets^10^ were *HSPB7, BAG3, CCT8* and *SVIL*. The former 3 loci were associated with HCM at P<5×10^−8^ while the locus mapped to *SVIL* did not reach GWAS significance (P=4.4×10^−6^ in HCM; P=2.9×10^−5^ in DCM; **Figure 2A-B**) and required further evidence to support implication in HCM. *SVIL* encodes supervillin, a large, multi-domain actin and myosin binding protein with multiple muscle and non-muscle isoforms, of which the muscle isoform has known roles in myofibril assembly and Z-disk attachment.^11^ SVIL is highly expressed in cardiac, skeletal, and smooth muscle myocytes in the Genotype Tissue Expression (GTEx) v9 single-nuclei RNA sequencing (snRNA-seq) dataset^12^, and *SVIL* morpholino knockdown in zebrafish produces cardiac abnormalities.^13^ In humans, loss of function (LoF) *SVIL* variants have been associated with smaller descending aortic diameter^14^ and homozygous LoF *SVIL* variants have been shown to cause a skeletal myopathy with mild cardiac features (left ventricular hypertrophy).^15^ To provide further evidence linking *SVIL* to human HCM and to explore the association of *SVIL* LoF variants with HCM, we performed rare variant burden analysis including 1,845 clinically-diagnosed unrelated HCM cases and 37,481 controls. We demonstrate a 10.5-fold (95% CI: 4.1-26.8; P:2.3×10^−7^) excess burden of *SVIL* LoF variants in HCM cases (**Figure 2C-D**; List of annotated variants in **Supplementary Table 6a**). Notably, the excess burden is even greater at 15.3-fold (95% CI: 5.7-41.3; P:7×10^−7^) when restricting the analysis to high confidence LoF variants affecting the predominant SVIL transcript in LV (ENST00000375400) (**Supplementary Table 6b**). In one family, the *SVIL* LoF variant (p.(Gln255*)) was carried by two cousins with HCM (parents deceased), providing some evidence of co-segregation. Taken together, these data support *SVIL* as a novel HCM disease gene.

**Figure 2:**
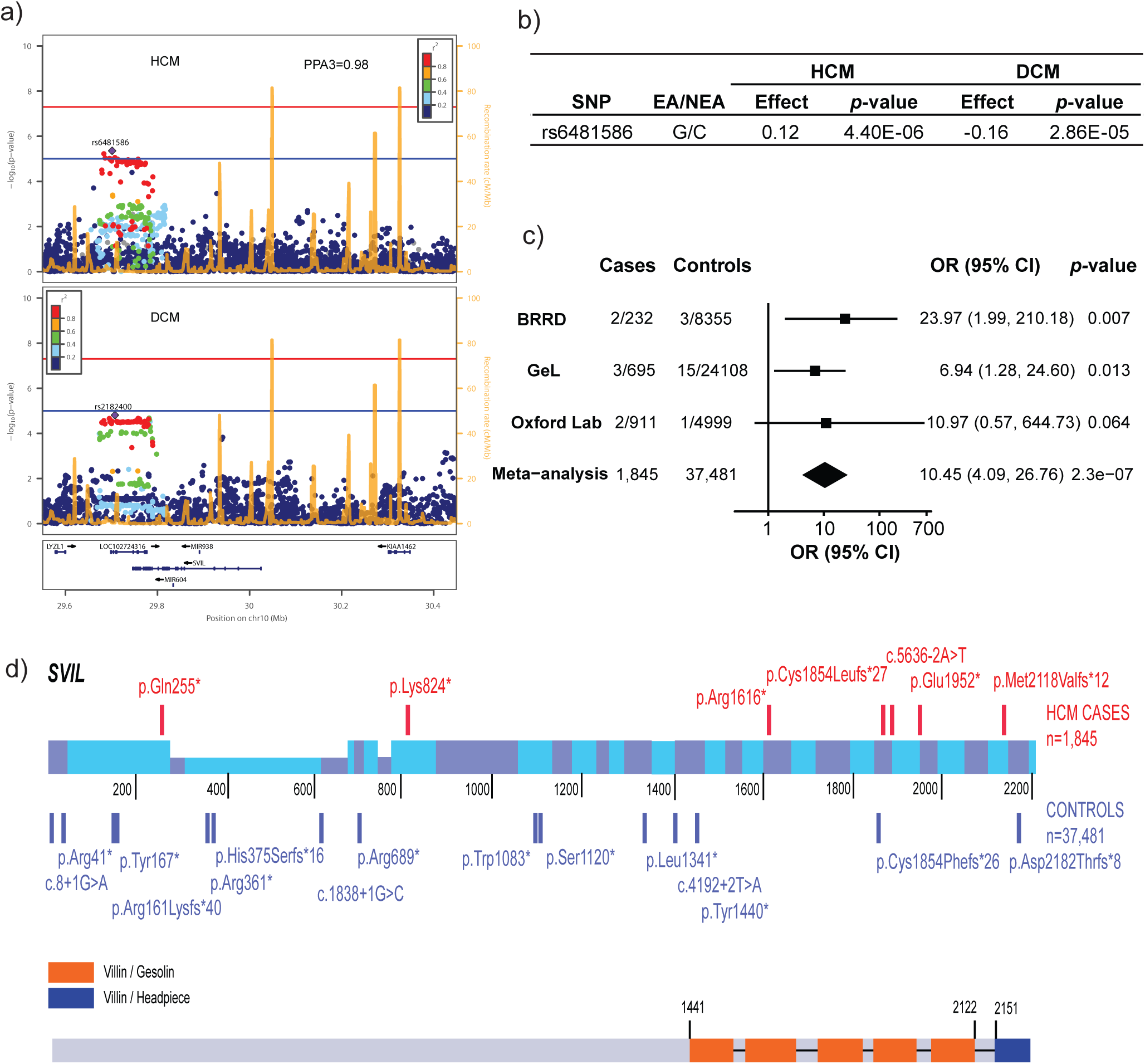
GWAS and rare variant association analyses identify *SVIL* as a novel HCM gene. **A**) GWAS in HCM and DCM^9^ identify a subthreshold locus near *SVIL*. GWAS-PW analysis identifies this locus as sharing the same causal variant (model 3) in both DCM and HCM (posterior probability of model 3, PPA3, 0.98). **B**) Summary statistics of the lead HCM variant (rs6481586) showing effect and non-effect alleles (EA/NEA) and opposite directions of effect (regression coefficient) in HCM and DCM. **C**) Forest plot showing excess of rare loss of function (LoF) variants in *SVIL* in HCM vs. controls in the Rare Disease Bioresource (BRRD), Genomics England (GeL) and Oxford laboratory. **D**) Schematic of the rare LoF *SVIL* variants in HCM cases (top, total N=1,845) and controls (bottom, total N=37,481) along the linear structure of SVIL. The coordinates reflect the codon numbers, and the coloured bars are the exons. The height of the exons reflects expression in cardiac isoforms and is not to scale. Detailed variant annotation appears in Supplementary Table 6.

To further maximize locus discovery in HCM, we performed a multi-trait analysis of GWAS (MTAG^16^; **Figure 3**). We first completed a GWAS of 10 cardiomyopathy-relevant LV traits in 36,083 participants of the UKB without cardiomyopathy and with available CMR, with machine learning assessment^17^ of LV volumes, wall thickness (mean and maximal) and myocardial strain (**Supplementary Table 7**; **Supplementary Figures 2-11**). We discovered 62 loci associated with LV traits (32 novel), of which 30 showed association with HCM with nominal significance (P<0.001) and 13 were mapped to genes associated with Mendelian heart disease (**Supplementary Table 8**). LDSC analyses^18^ demonstrated high genetic correlations (rg) between LV traits within 3 clusters (contractility, volume and mass) and with HCM (**Figure 3, Supplementary Table 9**). Leveraging such correlations, we then performed MTAG with HCM and 3 LV traits including the most correlated trait with HCM from each cluster, namely global circumferential strain (contractility cluster; rg -0.62), LV end-systolic volume (volume cluster; rg -0.48), and the ratio of LV mass to end-diastolic volume (mass cluster; rg 0.63). MTAG resulted in a significant increase in mean *χ*^2^ equivalent to ∼29% increase in effective sample size of the HCM GWAS (from 21,725 to 28,106), with an estimated upper bound of the false discovery rate (maxFDR)^16^ of 0.027. MTAG resulted in a substantial step up in loci discovered, identifying a total of 68 loci associated with HCM at P<5×10^−8^, including 48 that have not been previously published (13 novel ones also identified in the single-trait HCM meta-analysis, and 35 were additionally novel by MTAG) (**Figure 4, Supplementary Table 10**). Two of the 34 loci reaching genome-wide significance in the HCM GWAS were not significant in MTAG (loci mapped to *TRDN*/*HEY2* and *CHPF*). The total number of loci identified in GWAS or MTAG is therefore 70, of which 50 have not been previously published. Notably, the locus mapped to *SVIL* which was uncovered from the GWAS-PW analysis reached genome-wide significance in MTAG (P=1.1×10^−8^). Although it was not possible to test for replication for the 35 novel MTAG loci, a prior study strongly supports the robustness of the HCM-LV traits MTAG approach.^3^

**Figure 3:**
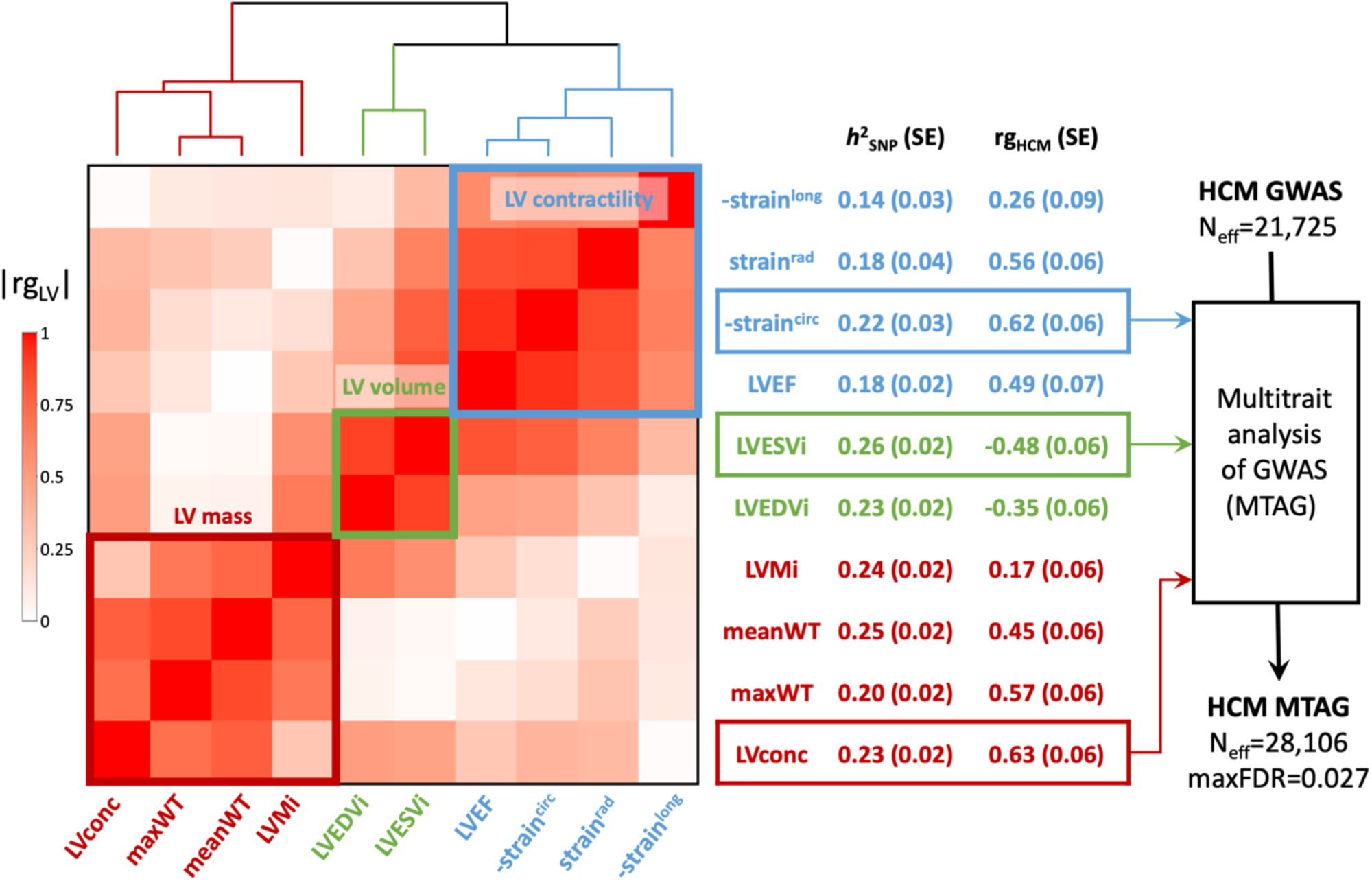
LV traits and HCM genetic correlations and use of MTAG to empower locus discovery. Pairwise genetic correlation between left ventricular (LV) traits shown in heatmap as absolute values (|rg_LV_|) ranging from 0 (white) to 1 (red). LV traits are sorted based on |rg_LV_| along the x and y axes using Euclidean distance and complete hierarchical clustering into 3 clusters: LV contractility (blue), volume (green) and mass (dark red). See dendrogram on top. The table in the middle shows the individual LV trait common variant heritability (*h*^2^_SNP_) and genetic correlation with HCM (rg_HCM_), with corresponding standard errors (SE). The trait with the strongest correlation (based on rg_HCM_) in each of the 3 clusters was carried forward for multi-trait analysis of GWAS (MTAG) to empower locus discovery in HCM. MTAG resulted in an increase of the effective sample size (N_eff_, based on number of cases and controls and increase in mean *χ*^2^ statistic) from 21,816 to 28,224, with an estimated upper bound of the false discovery rate (maxFDR) of 0.027. <u>Other abbreviations</u>: LVconc, LV concentricity index (LVM/LVEDV); LVEDVi, LV end-diastolic volume indexed for body surface area; LVEF, LV ejection fraction; LVESVi, LV end-systolic volume indexed for body surface area; LVMi, LV mass indexed for body surface area; maxWT, maximal LV wall thickness; meanWT, mean LV wall thickness; strain_circ_, global LV circumferential strain; strain_long_, global LV longitudinal strain; strain_rad_, global LV radial strain. <u>Note</u>: Since strain_circ_ and strain_long_ are negative values where increasingly negative values reflect increased contractility, we show -strain^circ^ and -strain^long^ to facilitate interpretation rg_HCM_ sign. Full rg_LV_ and rg_HCM_ results are shown in Supplementary Table 9.

**Figure 4:**
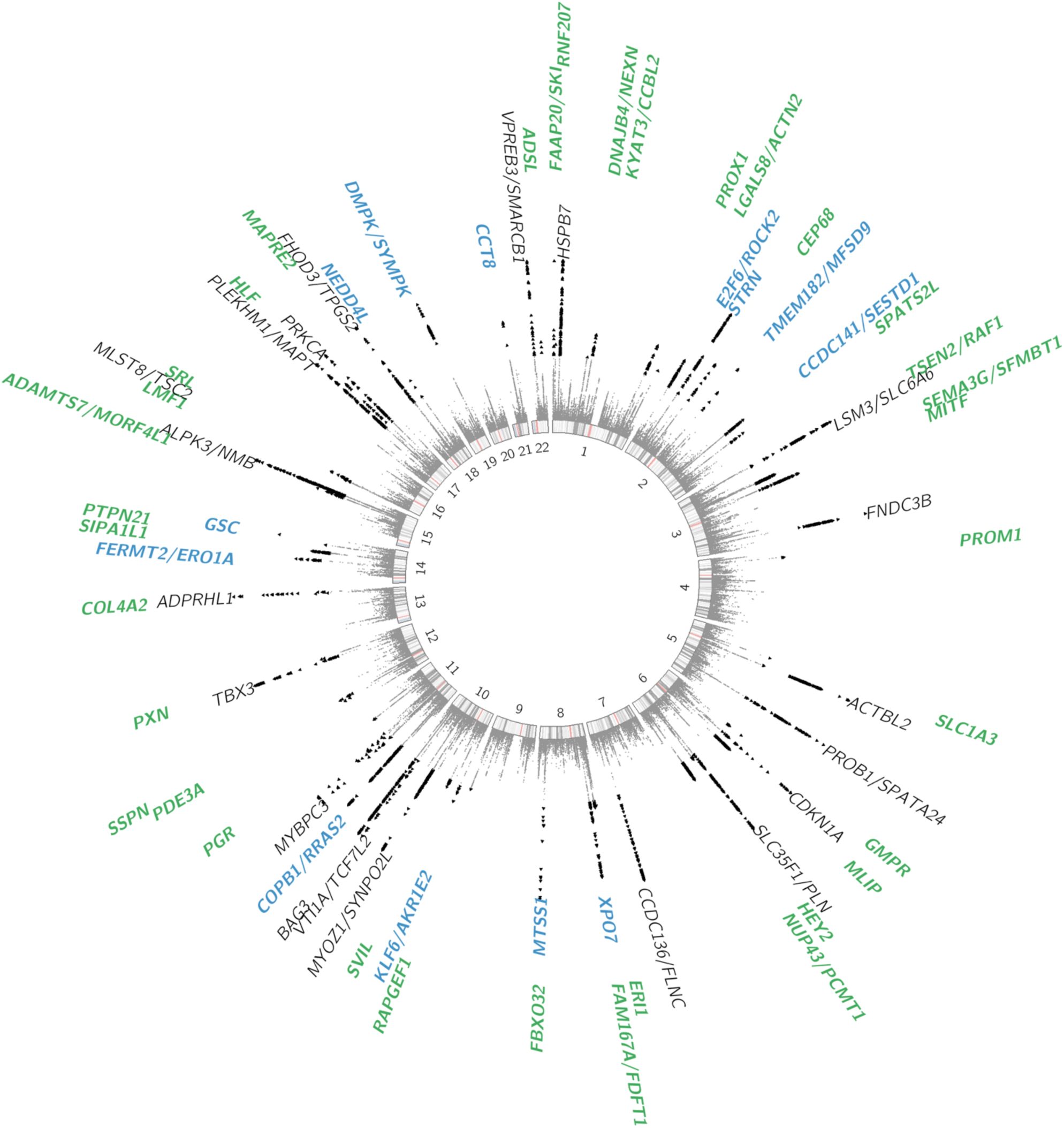
Circular Manhattan plot of HCM summary statistics from MTAG analysis. Previously published loci are identified in black (N=20), novel loci discovered by single trait all-comer GWAS meta-analysis are identified in blue (N=13) and additional novel loci from MTAG are identified in green (N=35). Two other loci reaching GWAS significance threshold in the single trait HCM GWAS meta-analysis but not reaching significance in MTAG are not shown (mapped to *TRDN*/*HEY2* and *CHPF*, see Table 1). Results with P<1×10^−15^ are assigned P=1×10^−15^. Variants with P<5×10^−8^ are shown as black triangles. Locus naming was performed primarily by OpenTargets gene prioritisation considering FUMA and prior gene association with Mendelian HCM. See Supplementary Table 10 for loci details.

MAGMA^19^ gene-set analysis identified multiple significant gene sets linked to muscle, contractility and sarcomeric function (**Supplementary Table 11**) and tissue expression analysis pointed to cardiac tissue (LV and atrial appendage, AA), and to a lower degree, other tissues with smooth muscle content, including arterial tissues (**Supplementary Table 12**). Within cardiac tissue, we further explored the contribution of specific cell types in HCM by leveraging available snRNA-seq data from donor human hearts.^20^ Using sc-linker^21^, we identified significant enrichment of heritability in cardiomyocyte and adipocyte cell types (cardiomyocyte: FDR-adjusted P=1.8×10^−6^; adipocyte: FDR-adjusted P=3.0×10^−3^) and state gene programs (**Supplementary Figure 12**).

Prioritization of potential causal genes in HCM MTAG loci was performed using OpenTargets variant to gene (V2G) mapping^10^ (**Supplementary Table 13**) and FUMA^22^ (**Supplementary Table 14**). Of all prioritized genes, 26 were selected based on concordance in both OpenTargets (top 3 genes per locus) and FUMA, as well as LV specific expression in bulk RNAseq data (GTEx v8) and expression in cardiomyocytes using publicly available snRNA-seq data from a recent study^23^ (**Supplementary Figure 13** and **Supplementary Tables 13-14**). Of those, 7 are known Mendelian cardiomyopathy genes (*PLN, FLNC, FHOD3* and *ALPK3* were previously reported^2,3^, while *ACTN2, TTN* and *NEXN* are in novel common variant HCM loci). Among the other 19 predominantly LV-expressed genes, 5 are mapped to previously published known HCM loci, while 14 are in novel loci and include genes involved in cardiomyocyte energetics and metabolism (*RNF207*^24^, *MLIP*^25^), myocyte differentiation and transcriptional regulation (*MITF*^26^, *PROX1*^27^, *TMEM182*^28^), myofibril assembly (*SVIL*^*11*^), and calcium handling and contractility (*PDE3A*^29^, *SRL*^30^). Last, a transcriptome-wide association study (TWAS) with S-MultiXcan^31^ using the MTAG summary statistics with cardiac tissues (LV and AA) from GTEx V8 identified 127 genes significantly associated with HCM at P<3.7×10^−6^ (**Supplementary Table 15**). Of those, 50 were not mapped to MTAG loci using either FUMA or OpenTargets, including *HHATL* (P=1×10^−11^), a gene of uncertain function prioritized based on dominant LV expression, and whose depletion in zebrafish may lead to cardiac hypertrophy.^32^

Rare sarcomeric variants associated with HCM have been shown to result in increased contractility, and cardiac myosin-inhibitors attenuate the development of sarcomeric HCM in animal models.^33^ Prior data from GWAS and Mendelian randomization (MR) also support a causal association of increased LV contractility with HCM, extending beyond rare sarcomeric variants.^3^ Pharmacologic modulation of LV contractility using myosin inhibitors has recently been approved in the treatment of HCM associated with LV obstruction (oHCM)^34,35^, but remains of uncertain utility in non-obstructive HCM (nHCM) which represents a significant proportion of the HCM patient population (both HCM_SARC-_ and HCM_SARC+_) and where no specific therapy currently exists. To further dissect the specific implication of LV contractility in nHCM and oHCM, we performed two-sample MR, testing the causal association of LV contractility as exposure, with HCM, nHCM and oHCM as outcomes. LV contractility was assessed with CMR using a volumetric method (LV ejection fraction, LVEF), and tridimensional tissue deformation methods (i.e. global LV strain in the longitudinal (strain_long_), circumferential (strain_circ_) and radial (strain_rad_) directions). Results from the primary MR inverse variance weighted (IVW) analysis are shown in **Figure 5A** and sensitivity analyses results appear in **Supplementary Table 16** and **Supplementary Figures 14-15**. Although significant heterogeneity in the exposure–outcome effects are limitations, MR findings support a causal association between increased LV contractility and increased risk for both nHCM and oHCM, with a substantial risk increase of 12-fold and 29-fold per standard deviation increase in strain_circ_, respectively (**Figure 5A**). Altogether, these data suggest that increased contractility is involved in both oHCM and nHCM development, and thus myosin inhibitors currently approved for symptom control in oHCM may also be of clinical benefit in nHCM. Last, we also performed MR analyses exploring whether increased systolic (SBP) and diastolic (DBP) blood pressure, and pulse pressure (PP=SBP-DBP) are causally associated with nHCM and oHCM. As for LV contractility, the causal association of SBP and DBP with HCM^2^ extended to both oHCM and nHCM subgroups (**Figure 5B, Supplementary Table 16** and **Supplementary Figure 16**), suggesting that lowering blood pressure may be a therapeutic target to mitigate disease progression for both nHCM and oHCM.

**Figure 5:**
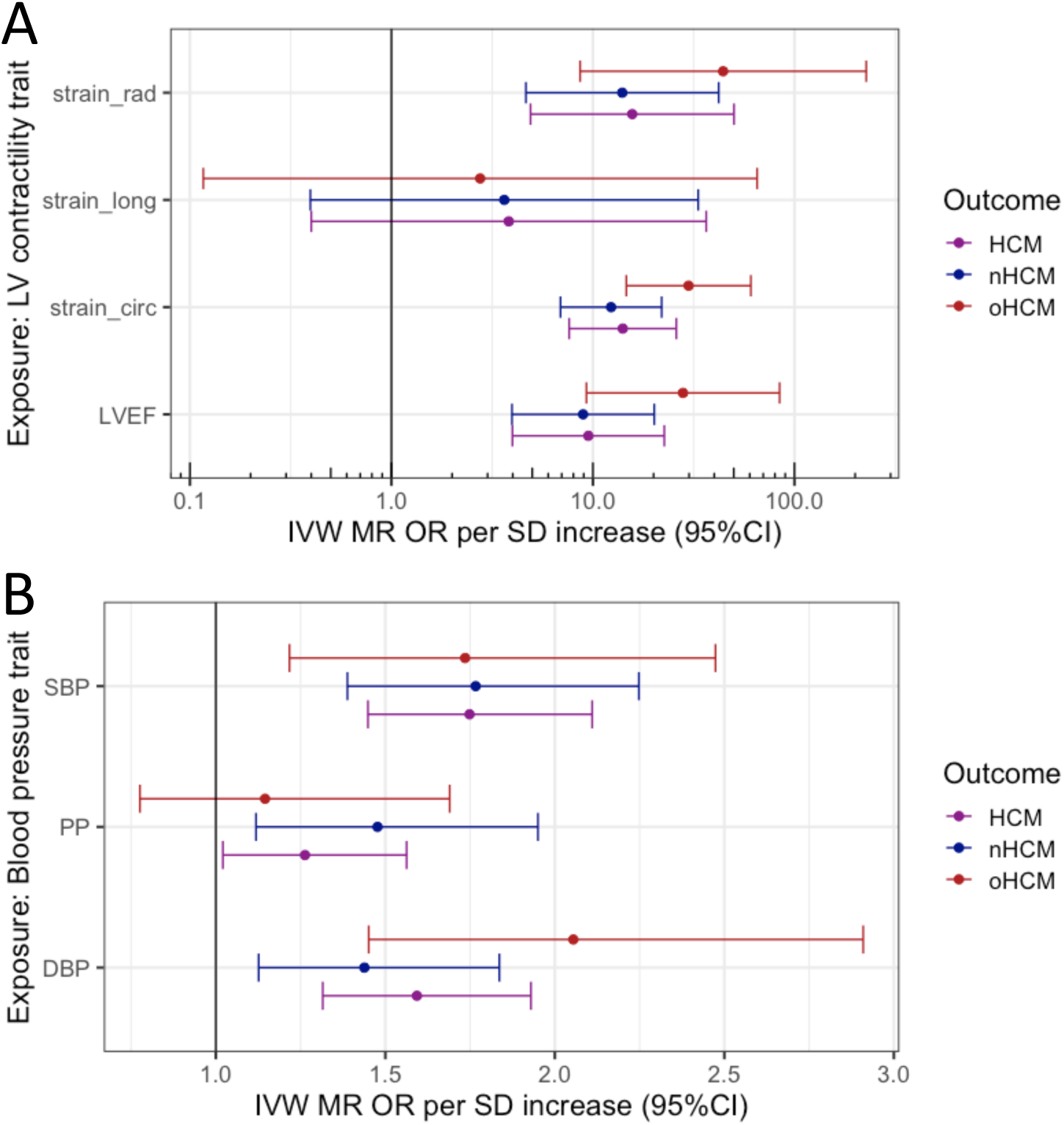
Mendelian randomization (MR) analysis of LV contractility and blood pressure on risk of obstructive (oHCM) and non-obstructive (nHCM) hypertrophic cardiomyopathy (HCM). Odds ratio (OR) represented are those inferred from the inverse variance weighted (IVW) two-sample MR per standard deviation increase (SD). The error bars represent the 95% confidence interval of the OR. **A)** MR suggests causal association of LV contractility (exposure) with HCM, oHCM and nHCM (outcomes), where increased contractility increases disease risk. Genetic instruments for LV contractility were selected from the present GWAS of left ventricular ejection fraction (LVEF), and strain in the radial (stain_rad), longitudinal (stain_long) and circumferential (strain_circ) directions in 36,083 participants of the UKB without cardiomyopathy and with available CMR. To facilitate interpretation of effect directions, OR for strain_circ and strain_long reflect those of increased contractility (more negative strain_circ and strain_long values). The outcome HCM GWAS included 5,927 HCM cases vs. 68,359 controls. Of those, 964 cases and 27,163 controls were included in the oHCM GWAS, and 2,491 cases and 27,109 were included in the nHCM GWAS. Note a logarithmic scale in the x-axis. **B)** MR suggests causal associations of systolic (SBP) and diastolic (DBP) blood pressure with HCM, nHCM and oHCM. Genetic instruments for SBP, DBP and pulse pressure (PP = SBP-DBP) were selected from a published GWAS including up to 801,644 individuals.^36^ See Supplementary Table 16 for full MR results.

In conclusion, the large number of new susceptibility loci arising from this work support new inferences regarding disease mechanisms in HCM. With the identification of the role of *SVIL*, we have uncovered further evidence that a subset of genes underlies both monogenic and polygenic forms of the condition. However, this shared genetic architecture does not extend to the core sarcomere genes which cause monogenic HCM; instead, the common variant loci implicate processes outside the myofilament, thereby widening our biological understanding and pointing to the importance of downstream remodeling pathways. These insights have therapeutic implications. The shared mechanistic pathways between obstructive and non-obstructive forms of HCM suggest that the new class of myosin inhibitors may be effective in both settings, while the further exploration of newly implicated loci and pathways may in the future yield new treatment targets.

## Methods

### GWAS of hypertrophic cardiomyopathy

The HCM GWAS included HCM cases and controls from 7 strata: the Hypertrophic Cardiomyopathy Registry (HCMR), a Canadian HCM cohort, a Netherlands HCM cohort, the Genomics England 100K Genome Project (GEL), the Royal Brompton HCM cohort, an Italian HCM cohort and the BioResource for Rare Disease (BRRD) project. Quality control (QC) and association analyses were performed per strata, followed by a meta-analysis. The 7 strata are described in the **Supplementary Note** and in **Supplementary Table 1**. Cases consisted of unrelated patients diagnosed with HCM in presence of unexplained left ventricular (LV) hypertrophy defined as a LV wall thickness (LVWT) >15mm, or >13mm and either presence of family history of HCM or a pathogenic or likely pathogenic genetic variant causing HCM. HCM cases underwent gene panel sequencing as per clinical indications. Variants identified within 8 core sarcomere genes (*MYBPC3, MYH7, TNNI3, TNNT2, MYL2, MYL3, ACTC1* and *TPM1*) were centrally assessed at the Oxford laboratory using the American College of Medical Genetics and Genomics (ACMG) guidelines.^38^ HCM cases were dichotomised into sarcomere-positive and sarcomere-negative groups using a classification framework previously reported in Neubauer et al.^39^ In addition to the primary all-comer GWAS analyses including all cases with HCM (total of 5,900 cases and 68,359 controls), analyses stratified for sarcomere status in cases and randomly allocated controls were performed, including a total of 1,776 cases vs. 29,414 controls in the sarcomere-positive analysis (HCM_SARC+_) and 3,860 cases vs. 38,942 controls in the sarcomere-negative analysis (HCM_SARC-_).

Meta-analyses for the all-comer HCM GWAS was performed on betas and standard errors using GWAMA.^40^ We kept variants where meta-analysis came from 2 or more studies and also had a sample size >5,000. Genomic inflation was estimated from the median *χ*^2^ distribution and using HapMap3 European ancestry LD scores using LD Score Regression.^5^ All variants were mapped to Genome Reference Consortium Human Build 37 (GRCh37) extrapolated using the 1000 Genome phase 3 genetic maps. A genome wide significant locus was assigned where two variants had a meta-analysis P<5×10^−8^ and were 0.5 cM distance apart. A similar approach was implemented for the HCM_SARC+_ and HCM_SARC-_ stratified analyses which comprised 5 and 7 strata, respectively (the GEL and BRRD strata did not include enough sarcomere-positive HCM cases). Variants were retained where meta-analysis came from 2 or more studies and had sample size >5,000 for sarcomere-negative and >2,500 for sarcomere-positive. The final dataset included 9,492,702 (all comer), 7,614,734 (HCM_SARC+_) and 9,226,079 (HCM_SARC-_) variants after filtering. The results of the all-comer HCM GWAS meta-analysis and stratified analyses are presented in **Table 1, Supplementary Figure 1** and **Supplementary Table 2**.

A false discovery rate (FDR) 1% P value cut-off was derived from the all-comer, HCM_SARC+_ and HCM_SARC-_ summary statistics using Simes method (Stata 10.1) and the corresponding P-values were 8.5×10^−6^, 1.6×10^−6^ and 7.8×10^−6^ respectively. Using the 1% FDR P value thresholds, we then performed a stepwise model selection to identify 1% FDR independently associated variants using GCTA.^4^ The analysis was performed chromosome wise using default window of 10Mbp, 0.9 collinearity and UKB reference panel containing 60K unrelated European ancestry participants. The results of this conditional analysis are presented in **Supplementary Table 3**.

### HCM heritability attributable to common variants

We estimated the heritability of HCM attributable to common genetic variation (*h*^2^_SNP_) in the all-comer HCM, as well as HCM_SARC+_ and HCM_SARC_ using LD score regression (LDSC)^5^ and genome-based restricted maximum likelihood (GREML)^6^. For LDSC, HapMap3 SNPs were selected from the summary statistics corresponding to HCM, HCM_SARC+_ and HCM_SARC_ meta-analyses. The *h*^2^_SNP_ was computed on the liability scale assuming a disease prevalence of 0.002.^41^ Since LDSC tends to underestimate *h*^2^_SNP_, we also estimated *h*^2^_SNP_ using GREML, as previously performed.^2,3^ We first computed *h*^2^_SNP_ for HCM, HCM_SARC+_ and HCM_SARC_, using GREML for each of the largest 3 strata (HCMR, the Canadian HCM cohort and the Netherlands HCM cohort), followed by fixed-effects and random-effects meta-analyses combining all 3 strata. To exclude the contribution of rare founder HCM causing variants, we excluded the *MYBPC3* locus for the Canadian and Netherlands strata and the *TNNT2* locus for the Canadian stratum.^3^ The results of *h*^2^_SNP_ analyses are presented in **Supplementary Table 4**.

### Locus colocalization in dilated (DCM) and hypertrophic cardiomyopathy (HCM)

We explored colocalization of HCM and DCM loci using GWAS-PW.^8^ The genome was split into 1,754 approximately independent regions and the all-comer HCM meta-analysis results were analysed with those of a publicly available DCM GWAS^9^ using a Bayesian approach. GWAS-PW fits each locus into one of the 4 models where model 1 is association in only the first trait, model 2 is association in only the second trait, model 3 when the two traits co-localize and model 4 when the genetic signals are independent in the two traits. We considered a locus to show colocalization when either trait harbours a genetic signal with P<1×10^−5^ and the GWAS-PW analysis demonstrates a posterior probability of association for model 3 (PPA3) greater than 0.8. Results of GWAS-PW are presented in **Supplementary Table 5** and **Figure 2** (panels **A** and **B**, for the *SVIL* locus).

### Association of rare *SVIL* loss of function (LoF) variants with HCM

We assessed the association of LoF variants in *SVIL* with HCM in 3 cohorts (BRRD^42^, GEL^43^ and the Oxford laboratory) followed by a meta-analysis. For BRRD, HCM cases were probands within the bio-resource project HCM. Controls were all remaining individuals within the BRRD projects except for those within the GEL and GEL2 projects (the Genomics England pilot data), since there is overlap of individuals with the GEL analysis in these two projects. For GEL, HCM cases were probands with a primary disease of HCM. Controls were probands without any primary or secondary cardiovascular disease and without any primary and secondary congenital myopathy, since *SVIL* has previously been associated with myopathy.^15^ For the Oxford laboratory, cases were clinically diagnosed with HCM and referred for diagnostic panel testing. The control group for the Oxford analysis consisted of 5,000 individuals randomly selected from the UK Biobank (UKB), which were all white British and unrelated. They had normal LV volume and function and no clinical diagnosis of cardiomyopathy (HCM or DCM). Genetic variants were identified using next generation sequencing (whole-genome sequencing for BRRD and GEL, panel/exome sequencing for Oxford cases and UKB controls) and annotated using the Ensembl variant effect predictor (VEP).^44^ LoF variants in *SVIL* were defined as those with the following VEP terms: stop lost, stop gained, splice donor variant, splice acceptor variant and frameshift variant. Only variants with a MAF<10^−4^ in the non-Finish European ancestral group of gnomAD v2.1.1^45^ were selected. Only LoF variants present in the Matched Annotation from NCBI and EMBL-EBI (MANE) / canonical transcript (NM_021738.3; ENST00000355867.9) were retained for the analysis. The proportion of cases and controls with *SVIL* LoF variants were compared using the Fisher Exact test for each of the 3 case-control datasets, followed by a fixed-effect model meta-analysis. We also performed a secondary analysis where association of *SVIL* LoF variants with HCM was restricted to variants that cause LoF in the primary LV transcript (ENST00000375400), and excluding variants expected to escape nonsense-mediated decay. The results of *SVIL* LoF variant association with HCM are shown in **Figure 2C**, and the list of *SVIL* LoF variants identified in cases and controls is shown in **Figure 2D** with annotation in **Supplementary Table 6a**. Results of the secondary analysis restricted to high confidence LoF variants are shown in **Supplementary Table 6b**.

### GWAS of cardiac magnetic resonance-derived left ventricular traits

#### UK Biobank (UKB) study population

The UKB is an open-access population cohort resource that has recruited half a million participants in its initial recruitment phase, from 2006-2010. At the time of analysis, CMR imaging data was available from 39,559 individuals in the imaging substudy. The UKB CMR acquisition protocol has been described previously.^46^ In brief, images were acquired according to a basic cardiac imaging protocol using clinical 1.5 Tesla wide bore scanners (MAGNETOM Aera, Syngo Platform VD13A, Siemens Healthcare, Erlangen, Germany) in three separate imaging centers. Extensive clinical and questionnaire data and genotypes are available for these individuals. Clinical data were obtained at the time of the imaging visit. These included sex (31), age (21003), weight (21002), height (50), SBP (4080), DBP (4079), self-reported non-cancer illness code (20002), and ICD10 codes (41270). The mean age at the time of CMR was 63 ± 8 (range 45-80), and 46% of participants were male. Cohort anthropometrics, demographics and comorbidities are reported in **Supplementary Table 7**. Exclusion criteria for the UKB imaging substudy included childhood disease, pregnancy and contraindications to MRI scanning. For the current analysis, we also excluded, by ICD-10 code and/or self-reported diagnoses, any subjects with heart failure, cardiomyopathy, a previous myocardial infarction, or structural heart disease. After imaging quality control and exclusions for comorbidities or genotype quality control, we had a maximum cohort size of 36,083 individuals. The UKB received National Research Ethics Approval (REC reference 11/NW/0382). The present study was conducted under terms of UKB access approval 18545.

#### LV trait phenotyping

Description of CMR image analysis has previously been published^3^ and is detailed in the **Supplementary Note**. We included ten LV phenotypes for GWAS analyses: end-diastolic volume (LVEDV), end-systolic volume (LVESV), ejection fraction (LVEF), mass (LVM), concentricity index (LVconc = LVM/LVEDV), mean wall thickness (meanWT), maximum wall thickness (maxWT) as well as global peak strain in radial (strain_rad_), longitudinal (strain_long_) and circumferential (strain_circ_) directions. The means and standard deviations of all ten LV phenotypes, overall and stratified by sex, are shown in **Supplementary Table 7**.

#### LV trait genome-wide association analyses

A description of genotyping, imputation and QC appears in the **Supplementary Note**. The GWAS model for LVEF, LVconc, meanWT, maxWT, strain_rad_, strain_long_ and strain_circ_ included age, sex, mean arterial pressure (MAP), body surface area (BSA, derived from the Mosteller formula) and the first eight genotypic principal components as covariates. LVEDV, LVESV and LVM were indexed to body surface area for the analysis, as commonly performed in clinical practice. For indexed values (LVEDVi, LVESVi, LVMi), the GWAS model did not include BSA as a covariate, but all other covariates were the same as for non-indexed phenotypes. BOLT-LMM (v2.3.2)^47^ was used to construct mixed models for association with around 9.5 million directly genotyped and imputed SNPs. A high-quality set of directly genotyped model SNPs was selected to account for random effects in the genetic association analyses. These were selected by MAF (>0.001), and LD-pruned (*r*^2^ <0.8) to create an optimum SNP set size of around 500,000. The model was then applied to the > 9.8 million imputed SNPs passing quality control and filtering. Results of the LV traits GWAS are shown in **Supplementary Table 8** and **Supplementary Figures 2-11**.

#### Locus definition and annotation

Genomic loci associated with all LV traits were annotated jointly. Specifically, summary statistics were combined and a P value corresponding to the minimal P value (minP) across all 10 summary statistics. The minP summary statics was then used to define loci using FUMA v1.4.2^22^ using a maximum lead SNP P-value of 5×10^−8^, maximum GWAS P-value of 0.05 and r^2^ threshold for independent significant SNPs of 0.05 (using the European 1000 Genomes Project dataset), and merging LD blocks within 250kb. Loci were then mapped to genes using positional mapping (<10kb), eQTL mapping using GTEx v8 restricted to atrial appendage, left ventricle and skeletal muscle tissues, and chromatin interaction mapping using left and right ventricles. See FUMA tutorial for detailed methods. Genes mapped using FUMA were further prioritized by querying the Clinical Genomes Resource (ClinGen)^48^ for genes linked to Mendelian heart disease with moderate, strong or definitive evidence, and using a recent review of overlapping GWAS and Mendelian cardiomyopathy genes.^37^ In addition to FUMA locus to gene mapping, we also report closest gene and top gene mapped using OpenTargets.^10^ Annotated LV trait loci are shown in **Supplementary Table 8**.

### Genetic correlations between HCM and LV traits

Pairwise genetic correlations for HCM and the 10 LV traits were assessed using LD score regression (LDSC, v.1.0.1).^18^ The analysis was restricted to well-imputed non-ambiguous HapMap3 SNPs, excluding SNPs with MAF<0.01 and those with low sample size, using default parameters. We then assessed genetic correlations for each of the 55 pairs (HCM and 10 LV traits) using precomputed LD scores from the European 1000 Genomes Project dataset. We did not constrain the single-trait and cross-trait LD score regression intercepts. The results of the genetic correlation analyses are shown in **Figure 3** and **Supplementary Table 9**.

### Multitrait analysis of GWAS (MTAG)

We performed multi-trait analysis of GWAS summary statistics using MTAG (v.1.0.8)^16^ to increase power for discovery of genetic loci associated with HCM. MTAG jointly analyzes multiple sets of GWAS summary statistics of genetically correlated traits to enhance statistical power. Due to high computation needs to calculate the maximum false discovery rate (maxFDR) with MTAG, we limited the number of GWAS summary statistics to 4 (HCM + 3 LV traits). The 3 LV traits to include were selected as follows. First, we performed hierarchical clustering of the 10 LV traits using the absolute value of the pairwise genetic correlations, Euclidean distance and the complete method, predefining the number of clusters to 3. This resulted in clustering of LV traits into a LV contractility cluster (LVEF, strain_rad_, strain_long_ and strain_circ_), a LV volume cluster (LVEDVi, LVESVi) and a LV mass cluster (LVMi, LVconc, meanWT, maxWT) (**Figure 3**). We then selected the trait with the highest genetic correlation with HCM for each cluster (strain_circ_, LVESVi and LVconc) to include in MTAG together with HCM. Only SNPs included in all meta-analyses (that is HCM and LV traits) were used in MTAG. The coded/noncoded alleles were aligned for all 4 studies before MTAG, and multi-allelic SNPs were removed. All summary statistics refer to the positive strand of GRCh37 and, as such, ambiguous/palindromic SNPs (having alleles A/T or C/G) were not excluded. Regression coefficients (beta) and their standard errors were used as inputs for MTAG. The maxFDR was calculated as suggested by the MTAG developers.^16^ MaxFDR calculates the type I error in the analyzed dataset for the worst-case scenario. We estimated the gain in statistical power by the increment in the effective sample size (N_eff_). The N_eff_ for the HCM GWAS was calculated using the following formula.^16,49^

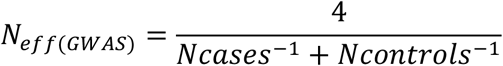

The N_eff_ for the HCM MTAG was estimated by means of the fold-increase in mean *χ*^2^, using the following formula.^16^

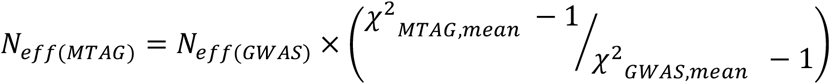

The MTAG N_eff_ corresponds to the approximate the sample size needed to achieve the same mean *χ*^2^ value in a standard GWAS. The results of HCM MTAG are presented in **Figure 4** and **Supplementary Table 10**.

### Genome-wide annotation

Genome-wide analyses following MTAG were performed using MAGMA v.1.08, as implemented in FUMA^22^, including gene-set and tissue expression analyses. We used Gene Ontology (GO) gene sets from the Molecular Signatures Database (MSigDB, v.6.2) for the gene-set analysis and the Genotype-Tissue Expression project (GTEx, v.8) for the tissue specificity analysis. The results of MAGMA analyses are shown in **Supplementary Table 11** (gene-set analyses) and **Supplementary Table 12** (tissue specificity analyses).

### Cardiac cell type heritability enrichment analysis

Gene programs derived from single nuclei RNA sequencing (snRNA-seq) were used to investigate heritability enrichment in cardiac cell types and states using the sc-linker framework.^21^ This approach uses snRNA-seq data to generate gene programs that characterize individual cell types and states. These programs are then linked to genomic regions and the SNPs that regulate them by incorporating Roadmap Enhancer-Gene Linking^50,51^ and Activity-by-Contact models^52,53^. Finally, the disease informativeness of resulting SNP annotations is tested using stratified LD score regression (S-LDSC)^54^ conditional on broad sets of annotations from the baseline-LD model.^55,56^ Cell type and state-specific gene programs were generated from snRNA-seq data of ventricular tissue from 12 control subjects, with cell type and state annotations made as part of a larger study of ∼880,000 nuclei (samples from 61 DCM and 12 control subjects.^20^ Cell states that may not represent true biological states (for example, technical doublets) were excluded from analysis. Results of sc-linker cardiac cell type heritability enrichment analysis are shown in **Supplementary Figure 12**.

### Locus to gene annotation

A genome wide significant HCM MTAG loci was assigned where two variants had a MTAG P<5×10^−8^ and were 0.5 cM distance apart, as performed for the HCM GWAS. Prioritization of potential causal genes in HCM MTAG loci was performed using OpenTargets variant to gene (V2G) mapping^10^ and FUMA^22^. The lead SNP at each independent locus was used as input for OpenTargets V2G using the release of October 12^th^, 2022. Locus to gene mapping with FUMA v1.3.7 was performed based on 1) position (within 100kb), 2) eQTL associations in disease-relevant tissues (GTEx V8 left ventricle, atrial appendage and skeletal muscle) and 3) chromatin interactions in cardiac tissue (left ventricle and right ventricle, FDR P<10^−6^).

We further annotated genes mapped using OpenTargets and/or FUMA with their implication in mendelian cardiomyopathy. Specifically, we queried the Clinical Genome Resource (ClinGen^48,57^) for genes associated with any cardiomyopathy phenotype with a level of evidence of moderate, strong or definitive and included genes with robust recent data supporting an association with Mendelian cardiomyopathy.^37^

We also prioritized genes based on RNA expression data from bulk tissue RNAseq data in the GTEx^58^ v8 dataset accessible at the GTEx portal and snRNA-seq data from Chaffin et al^23^ accessible through the Broad Institute single cell portal (singlecell.broadinstitute.org). Using the GTEx v8 data, we assessed specificity of LV expression by computing the ratio of median LV transcripts per million (TPM) to the median TPM in other tissues excluding atrial appendage and skeletal muscle and averaging tissue within types (e.g., all arterial tissues, all brain tissues, etc.). High and Mid LV expression specificity were empirically defined as >10-fold and >1.5-fold LV to other tissues median TPM ratios, respectively. Using snRNA-seq data from Chaffin et al^23^, we report the expression in the cardiomyocyte_1 cell type using scaled mean expression (relative to each gene’s expression across all cell types) and percentage of cells expressing. High and Mid expression in cardiomyocytes were empirically defined as percentage expressing cells ≥80% and 40-80%, respectively. Prioritized genes were defined as genes mapped using both OpenTargets (top 3 genes) and FUMA, AND had either 1) High LV specific expression, OR 2) High cardiomyocyte expression, OR 3) both Mid LV specific expression and Mid cardiomyocyte expression.

Gene mapping data including ClinGen cardiomyopathy genes at HCM loci, LV expression specificity and cardiomyocyte expression are shown in **Supplementary Table 13** (OpenTargets genes) and **Supplementary Table 14** (FUMA genes). Prioritized genes are illustrated in **Supplementary Figure 13**.

### Transcriptome-wide association study (TWAS)

We used MetaXcan to test the association between genetically predicted-gene expression and HCM using summary results from MTAG analysis.^31,59^ Biologically-informed MASHR-based prediction models of gene expression for left ventricle (LV) and atrial appendage (AA) tissue from GTEx v8^60^ were analysed individually with S-PrediXcan^59^, and then analysed together using S-MultiXcan.^31^ GWAS MTAG summary statistics were harmonised and imputed to match GTEx v8 reference variants present in the prediction model. To account for multiple testing, TWAS significance was adjusted for the total number of genes present in S-MultiXcan analysis (13,558 genes, P=3.7×10^−6^). TWAS results are shown in **Supplementary Table 15**.

### Two-sample Mendelian randomization (MR)

We assessed whether increased contractility and blood pressure are causally linked to increased risk of HCM globally and its obstructive (oHCM) and non-obstructive (nHCM) forms using two-sample MR. LV contractility and blood pressure parameters were used as exposure variables, and HCM, oHCM and nHCM as outcomes. Analyses were performed using the TwoSampleMR (MRbase) package^61^ (v.0.5.6) in R (v.4.2.0). Four exposure variables corresponding to measures of LV contractility were used separately: LVEF as a volumetric marker of contractility, and global strain (strain_circ_, strain_rad_ and strain_long_) as contractility markers based on myocardial tissue deformation. Instrument SNPs for contractility were selected based on the LV trait GWAS presented here using a P value threshold of <5×10^−8^. Only independent SNPs (using *r*^2^<0.01 in the European 1000 Genomes population) were included. Instrument SNPs for the blood pressure analysis were selected with a similar approach using a published blood pressure GWAS.^36^ The outcome summary statistics were those of the single-trait HCM case-control meta-analysis (5,927 cases and 68,359 controls). We also performed a GWAS meta-analysis including data from HCMR and the Canadian HCM cohort (**Supplementary Table 1**) for nHCM (2,491 cases and 27,109 controls) and oHCM (964 cases and 27,163 controls) to use as outcomes. For these stratified analyses, oHCM was defined as HCM in presence of a LV outflow tract gradient ≥30mmHg at rest or during Valsalva/exercise at any time point. All other HCM cases were considered nHCM. Notably, nHCM and oHCM show high genetic correlation (rg=0.87 with standard error, SE=0.13; P=4×10^−11^), suggesting a substantially shared genetic basis.

Insertions/deletions and palindromic SNPs with intermediate allele frequencies (MAF>0.42) were excluded, and other SNPs in the same locus were included only if P<5×10^−8^. An inverse variance weighted MR model was used as a primary analysis. We used three additional methods as sensitivity analyses: weighted median, weighted mode and MR Egger. Cochran’s Q statistics were calculated to investigate heterogeneity between SNP causal effects using IVW. Evidence of directional pleiotropy was also assessed using the MR Egger intercept. Mean F-statistics were calculated to assess the strength of the genetic instruments used. Leave-one-out analyses were also performed to ensure the SNP causal effects are not driven by a particular SNP. The summary results of MR analyses are shown in **Figure 5** and **Supplementary Table 16**, with effect plots shown in **Supplementary Figures 14** (contractility) and **Supplementary Figure 16** (blood pressure), and leave-one-out analyses for the contractility MR in **Supplementary Figure 15**. The MR effects are shown per unit change (% for contractility; mmHg for blood pressure) in **Supplementary Table 16** and **Supplementary Figures 14-16**, and per SD change in **Figure 5**. OR per SD increase are calculated as follows 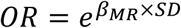. SDs are reported in **Supplementary Table 16** and correspond to those in the current UKB CMR dataset (for contractility) and those reported by Evangelou et al^36^ in the UK Biobank (for blood pressure).

## Data Availability

Data from the Genome Aggregation Database (gnomAD, v.2.1.1) are available at https://gnomad.broadinstitute.org. Data from the UKB can be requested from the UKB Access Management System (https://bbams.ndph.ox.ac.uk). Data from the GTEx consortium are available at the GTEx portal (https://gtexportal.org). Published snRNA-seq data are available at the Broad Single Cell Portal (singlecell.broadinstitute.org) and at the Cellxgene tool website (https://cellxgene.cziscience.com/collections/e75342a8-0f3b-4ec5-8ee1-245a23e0f7cb/private). Other datasets generated during and/or analyzed during the current study can be made available upon reasonable request to the corresponding authors. Individual level data sharing is subject to restrictions imposed by patient consent and local ethics review boards. Summary statistics of GWAS and MTAG will be made available in the GWAS catalog upon publication following peer-review, and interactive Manhattan and regional plots will be made available at www.well.ox.ac.uk/hcm.

## Code availability

The analyses reported in this manuscript rely on previously published software, as detailed in the methods section and in the reporting summary. Code of custom scripts will be made available upon request.

## Acknowledgements

This work was supported by funding from the British Heart Foundation (BHF, RG/18/9/33887, RE/18/4/34215, FS/15/81/31817); the National Heart, Lung, and Blood Institute (NIH grant U01HL117006-01A1); the Wellcome Trust (201543/B/16/Z, 107469/Z/15/Z, 200990/A/16/Z); the Wellcome Trust core awards (090532/Z/09/Z, 203141/Z/16/Z); the National Institute for Health Research (NIHR) Oxford Biomedical Research Centre; the NIHR Imperial College Biomedical Research Centre; NIHR Royal Brompton Cardiovascular Biomedical Research Unit; Sir Jules Thorn Charitable Trust [21JTA]; the Medical Research Council (MRC, UK); the Dutch Heart Foundation (CVON 2018-30 PREDICT2); the Horstingstuit Foundation; the Montreal Heart Institute Foundation; the Philippa and Marvin Carsley Cardiology Chair; the Fonds de la Recherche du Québec-Santé (254616, 265449); the Canadian Institutes for Health Research (CIHR, 428321). For the purpose of open access, the authors have applied a CC BY public copyright licence to any Author Accepted Manuscript version arising from this submission. The views expressed in this work are those of the authors and not necessarily those of the funders.

R.T. holds the Canada Research Chair in translational cardiovascular genetics. S.L.Z. received support from BHF Centre of Research Excellence Clinical Research Fellowship (RE/18/4/34215). C.F. received funding from a BHF Clinical Research Training Fellowship (FS/15/81/31817). A.R.H. received support from the MRC Doctoral Training Partnership. X.X. is currently a post-doc scientist funded by MRC-London Institute of Medical Sciences. R.W. received support from an Amsterdam Cardiovascular Sciences fellowship. M.T. receives support from Monat Foundation. S.J.J. was supported by a Junior Clinical Scientist Fellowship (03-007-2022-0035) from the Dutch Heart Foundation, and by an Amsterdam UMC Doctoral Fellowship. Y.M.P. receives support from the Dutch Heart Foundation (CVON PRIME). B.P.H. is funded by the BHF Intermediate Fellowship (FS/ICRF/21/26019). D.P.O’R. is supported by the MRC (MC_UP_1605/13) and BHF (RG/19/6/34387). R.A.d.B. is supported by the Dutch Heart Foundation (2020B005), by the Leducq Foundation (Cure-PLaN), and by the European Research Council (ERC CoG 818715). I.C. receives support from the Dutch Heart Foundation (CVON 2015-12 eDETECT). P.M.M. received funding from the Edmond J. Safra Foundation and Lily Safra and an NIHR Senior Investigator Award, the UK Dementia Research Institute, which receives its funding from UK DRI Ltd., funded by the MRC, Alzheimer’s Society, and Alzheimer’s Research UK and the Imperial College British Heart Foundation Centre of Excellence. J.-C.T. holds the Canada Research Chair in personalized medicine and the University of Montreal endowed research chair in atherosclerosis, and he is also the principal investigator of the Montreal Heart Institute André and France Desmarais hospital cohort funded by the Montreal Heart Institute Foundation. A.G. has received support from the BHF, European Commission [LSHM-CT-2007-037273, HEALTH-F2-2013-601456] and TriPartite Immunometabolism Consortium [TrIC]-NovoNordisk Foundation [NNF15CC0018486], BHF-DZHK (SP/19/2/344612). C.R.B. received support from EJP-RD (LQTS-NEXT, ZonMW project 40-46300-98-19009) and the Leducq Foundation (project 17CVD02). HW is member of the Oxford BHF Centre for Research Excellence (RE/13/1/30181). HW and JW are supported by CureHeart, the British Heart Foundation’s Big Beat Challenge award (BBC/F/21/220106).

This research has been conducted in part using the UK Biobank Resource under application numbers 18545 and 47602. This research was made possible through access to the data and findings generated by the 100,000 Genomes Project. The 100,000 Genomes Project is managed by Genomics England Limited (a wholly owned company of the Department of Health and Social Care), and funded by the National Institute for Health Research and NHS England. The Wellcome Trust, Cancer Research UK and the Medical Research Council have also funded research infrastructure. The 100,000 Genomes Project uses data provided by patients and collected by the National Health Service as part of their care and support. The Genotype-Tissue Expression (GTEx) Project was supported by the Common Fund of the Office of the Director of the National Institutes of Health, and by NCI, NHGRI, NHLBI, NIDA, NIMH, and NINDS. The data used for the analyses described in this manuscript were obtained from the GTEx Portal.

