## Supplemental Figures and Note for "Large scale genome-wide association analyses identify novel genetic loci and mechanisms in hypertrophic cardiomyopathy"


**Supplementary Figure 1**: Manhattan plots of HCM GWAS for all cases (A) and sarcomere positive/negative cases (B)

**Supplementary Figure 2**: Manhattan plot for GWAS of global left ventricular longitudinal strain

**Supplementary Figure 3**: Manhattan plot for GWAS of global left ventricular radial strain

**Supplementary Figure 4**: Manhattan plot for GWAS of global left ventricular circumferential strain

**Supplementary Figure 5**: Manhattan plot for GWAS of left ventricular ejection fraction

**Supplementary Figure 6**: Manhattan plot for GWAS of left ventricular end-systolic volume

**Supplementary Figure 7**: Manhattan plot for GWAS of left ventricular end-diastolic volume

**Supplementary Figure 8**: Manhattan plot for GWAS of left ventricular mass

**Supplementary Figure 9**: Manhattan plot for GWAS of mean left ventricular wall thickness

**Supplementary Figure 10**: Manhattan plot for GWAS of maximal left ventricular wall thickness

**Supplementary Figure 11**: Manhattan plot for GWAS of left ventricular concentricity index

**Supplementary Figure 12**: Enrichment of 68 cell-state (A) and 9 cell-type (B) gene programs

**Supplementary Figure 13**: HCM locus to gene mapping and prioritization based on cardiac expression

**Supplementary Figure 14**: SNP effects of contractility on risk of HCM, oHCM and nHCM

**Supplementary Figure 15**: Leave-one-out analyses for contractility Mendelian randomization

**Supplementary Figure 16**: SNP effects of blood pressure on risk of HCM, oHCM and nHCM

**Supplementary Note**

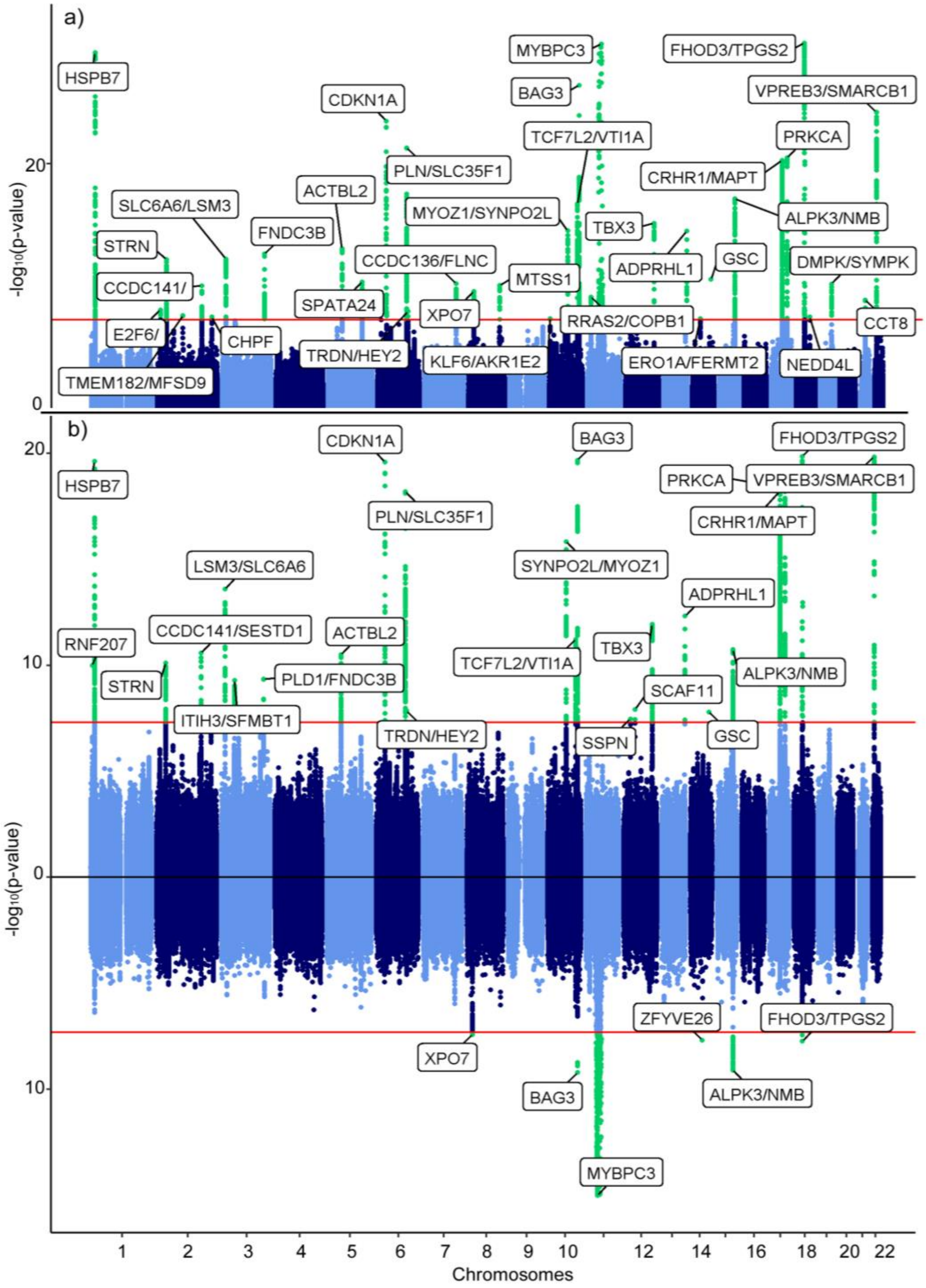


**Supplementary Figure 1: Manhattan plots of GWAS of HCM for all comers (A) and sarcomere positive/negative cases (B).** Sarcomere negative ($HCM_{SARC-}$) summary statistics are on the positive y-axis and sarcomere positive ($HCM_{SARC+}$) summary statistics on the negative y-axis in panel B. Red line denotes the significance threshold of $P=5x10^{-8}$. Plots are truncated at –log10(p-value) of 30 (a), 20 (S-) and 15 (S+). See Table 1 and Supplementary Table 2 for description of lead variants for the all-comer and stratified analyses ($HCM_{SARC-}$ and $HCM_{SARC+}$), respectively.

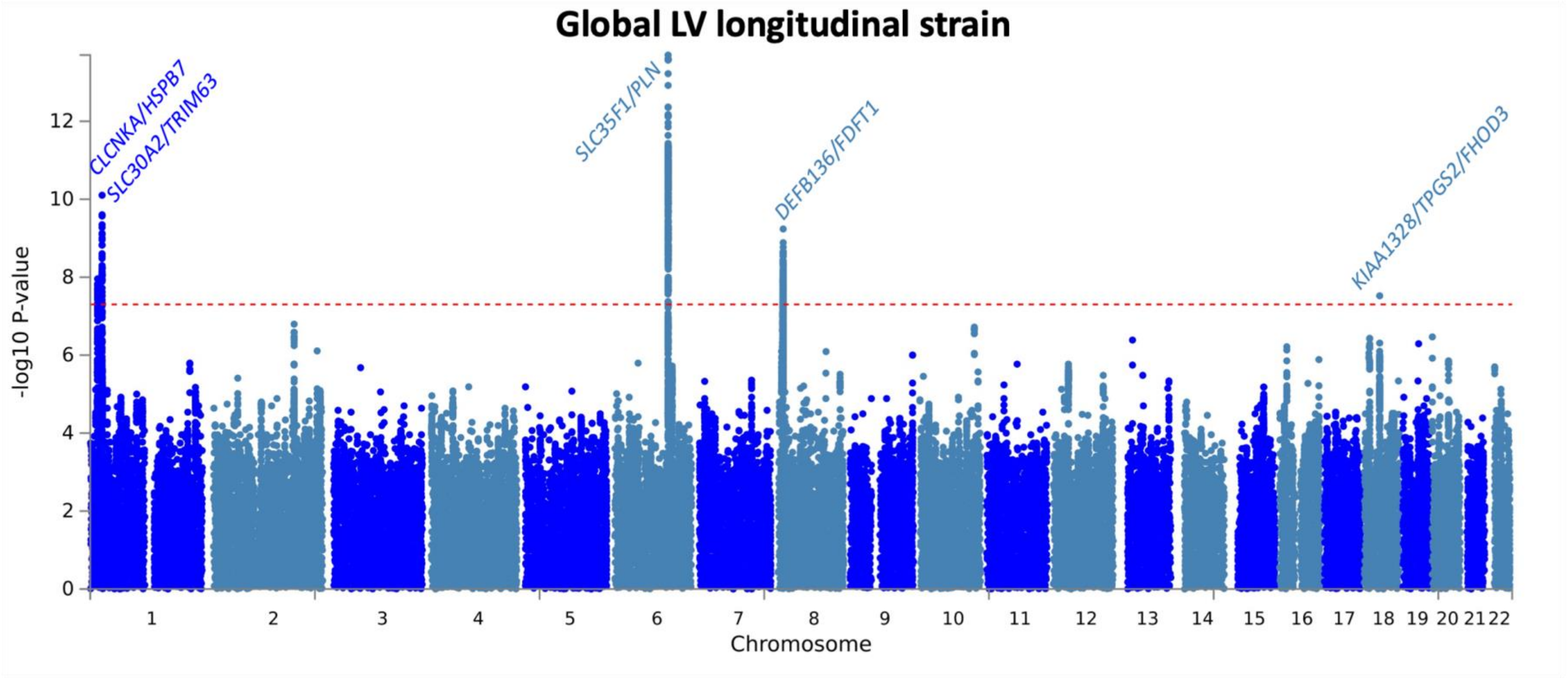


**Supplementary Figure 2:** Manhattan plot for GWAS of global left ventricular longitudinal strain. Based on an analysis of 35,052 UK Biobank participants without cardiomyopathy and with available cardiac magnetic resonance data. Genomic position presented on the x-axis and -log (P-value) on the y-axis. Manhattan plot generated using FUMA[1] and annotated with locus names based on nearest gene, top gene mapped using OpenTargets[2] variant to gene pipeline (25 February 2022 release), and any mendelian cardiomyopathy gene mapped using FUMA v1.3.8. Locus name and data point colors alternate by chromosome. See Supplementary Table 8 for description of the lead variants in the LV trait GWAS and lookup in the HCM GWAS.

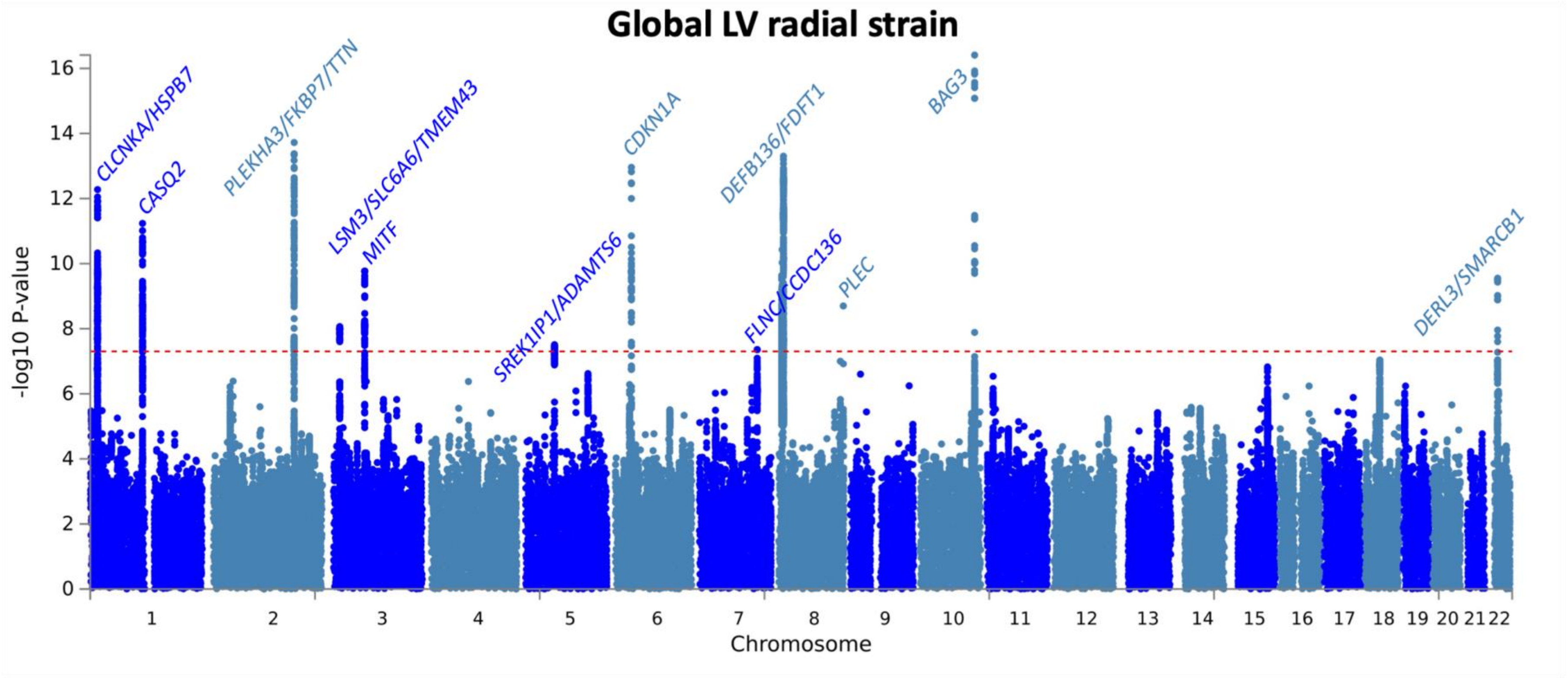


**Supplementary Figure 3:** Manhattan plot for GWAS of global left ventricular radial strain. Based on an analysis of 36,033 UK Biobank participants without cardiomyopathy and with available cardiac magnetic resonance data. Genomic position presented on the x-axis and -log (P-value) on the y-axis. Manhattan plot generated using FUMA[1] and annotated with locus names based on nearest gene, top gene mapped using OpenTargets[2] variant to gene pipeline (25 February 2022 release), and any mendelian cardiomyopathy gene mapped using FUMA v1.3.8. Locus name and data point colors alternate by chromosome. See Supplementary Table 8 for description of the lead variants in the LV trait GWAS and lookup in the HCM GWAS.

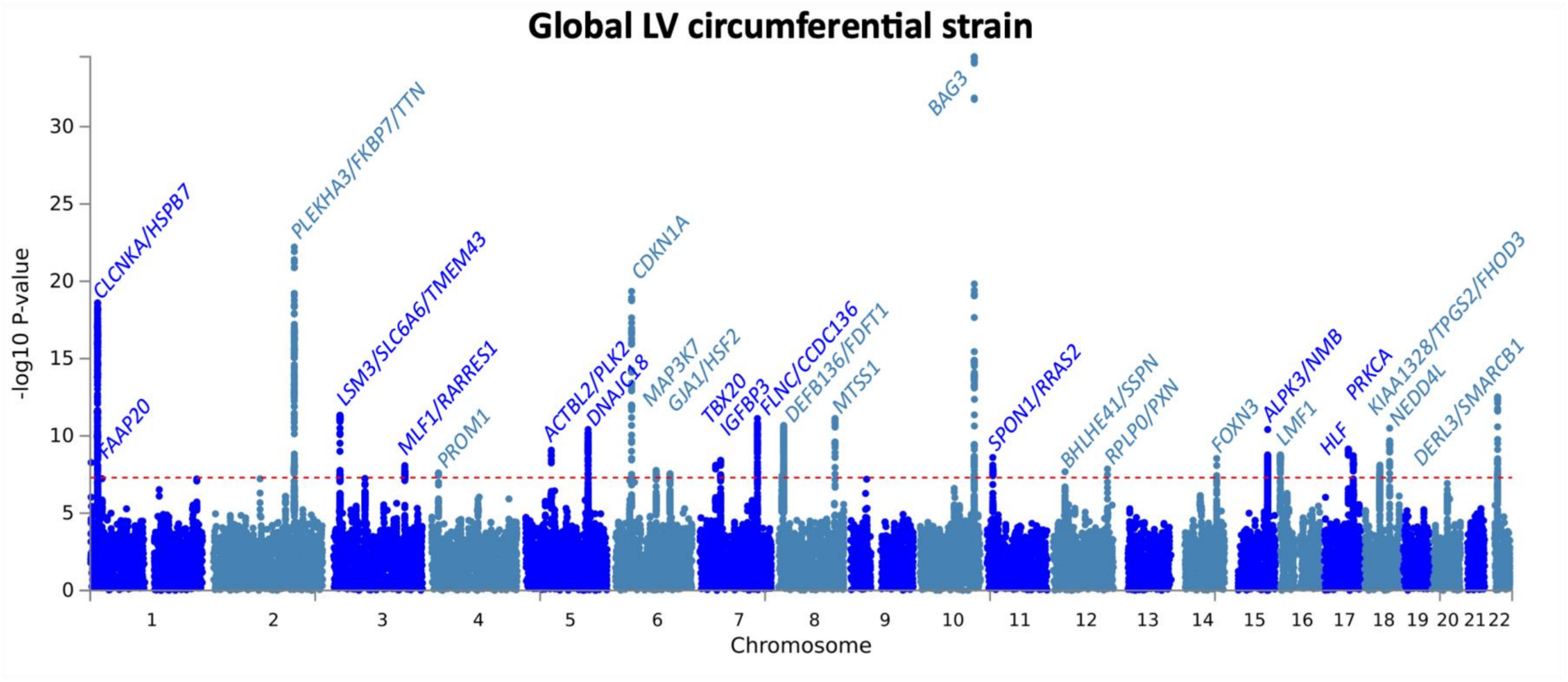


**Supplementary Figure 4:** Manhattan plot for GWAS of global left ventricular circumferential strain. Based on an analysis of 36,033 UK Biobank participants without cardiomyopathy and with available cardiac magnetic resonance data. Genomic position presented on the x-axis and -log (P-value) on the y-axis. Manhattan plot generated using FUMA[1] and annotated with locus names based on nearest gene, top gene mapped using OpenTargets[2] variant to gene pipeline (25 February 2022 release), and any mendelian cardiomyopathy gene mapped using FUMA v1.3.8. Locus name and data point colors alternate by chromosome. See Supplementary Table 8 for description of the lead variants in the LV trait GWAS and lookup in the HCM GWAS.

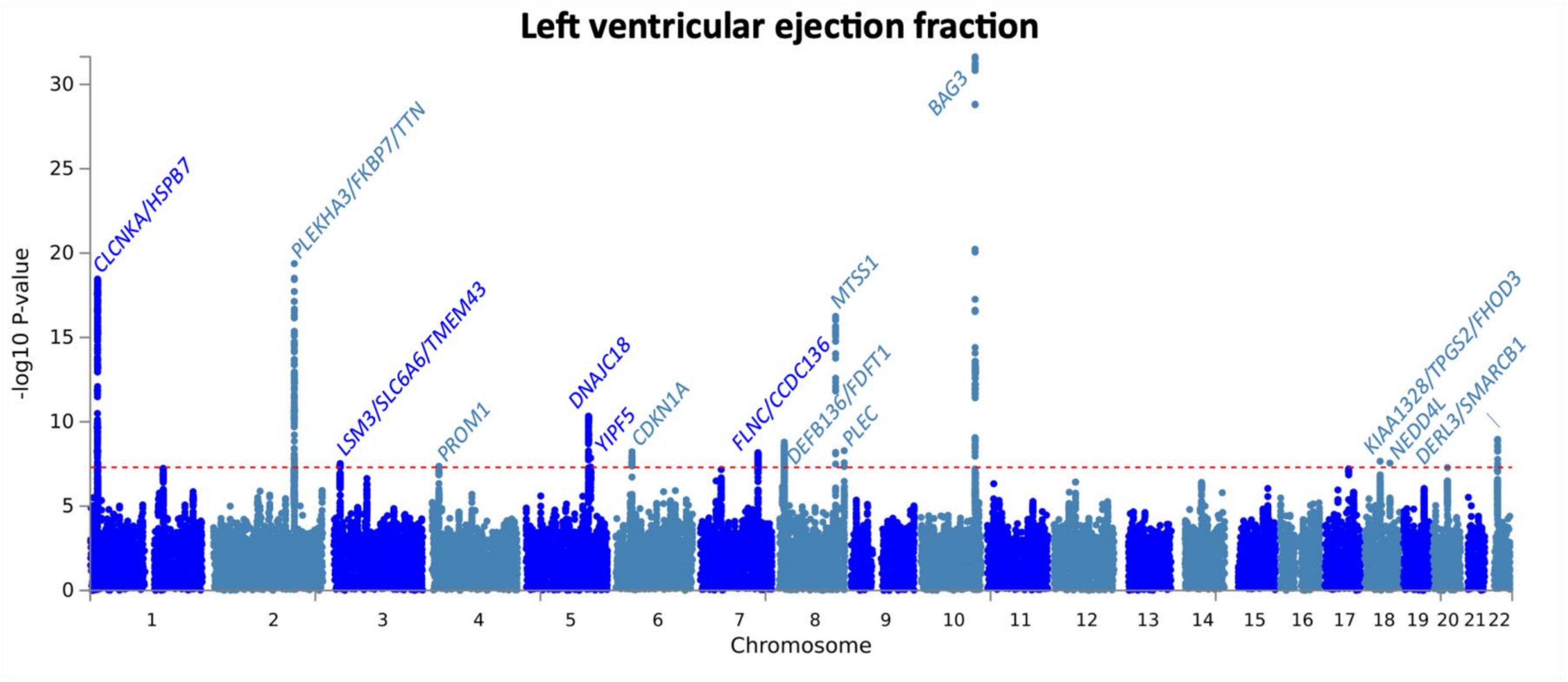


**Supplementary Figure 5:** Manhattan plot for GWAS of left ventricular ejection fraction. Based on an analysis of 36,083 UK Biobank participants without cardiomyopathy and with available cardiac magnetic resonance data. Genomic position presented on the x-axis and -log (P-value) on the y-axis. Manhattan plot generated using FUMA[1] and annotated with locus names based on nearest gene, top gene mapped using OpenTargets[2] variant to gene pipeline (25 February 2022 release), and any mendelian cardiomyopathy gene mapped using FUMA v1.3.8. Locus name and data point colors alternate by chromosome. See Supplementary Table 8 for description of the lead variants in the LV trait GWAS and lookup in the HCM GWAS.

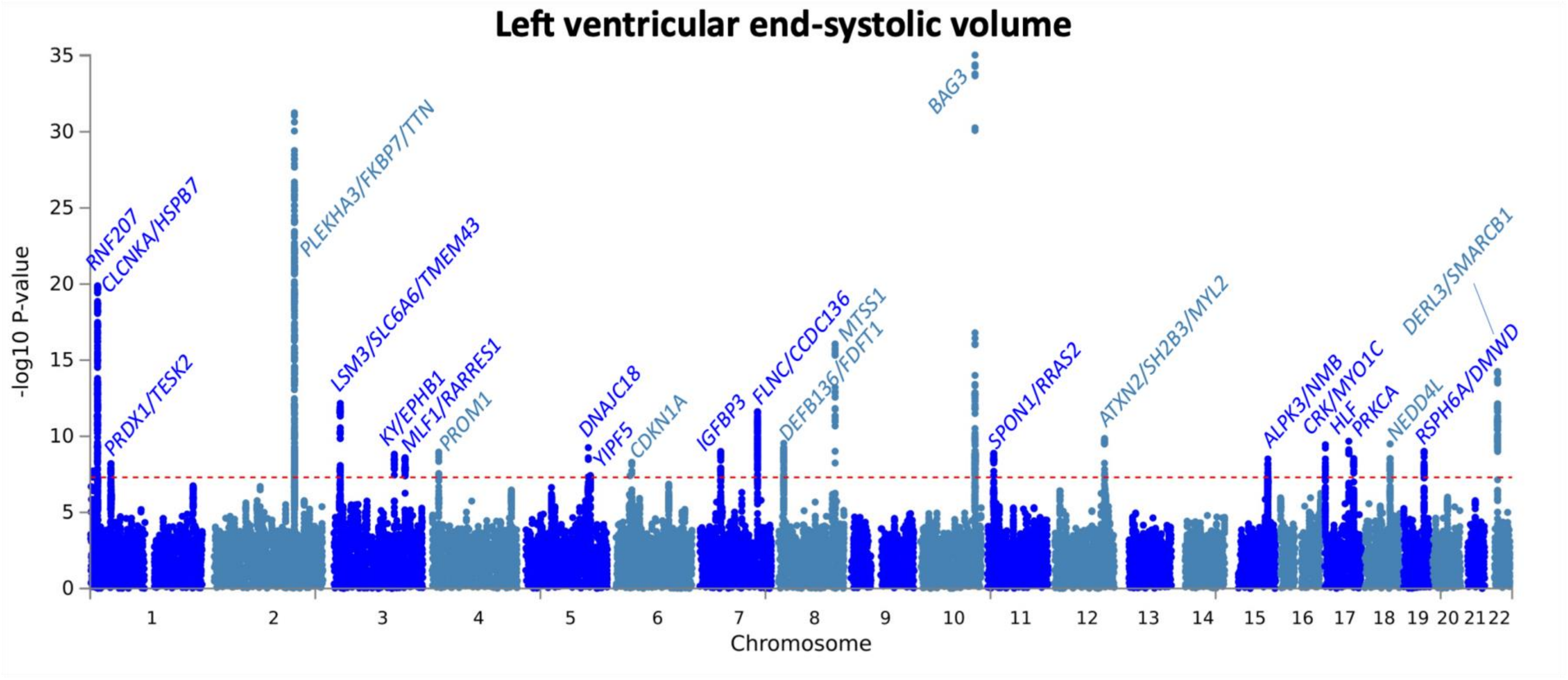


**Supplementary Figure 6:** Manhattan plot for GWAS of left ventricular end-systolic volume, indexed to the body surface area. GWAS based on an analysis of 36,083 UK Biobank participants without cardiomyopathy and with available cardiac magnetic resonance data. Genomic position presented on the x-axis and -log (P-value) on the y-axis. Manhattan plot generated using FUMA[1] and annotated with locus names based on nearest gene, top gene mapped using OpenTargets[2] variant to gene pipeline (25 February 2022 release), and any mendelian cardiomyopathy gene mapped using FUMA v1.3.8. Locus name and data point colors alternate by chromosome. See Supplementary Table 8 for description of the lead variants in the LV trait GWAS and lookup in the HCM GWAS.

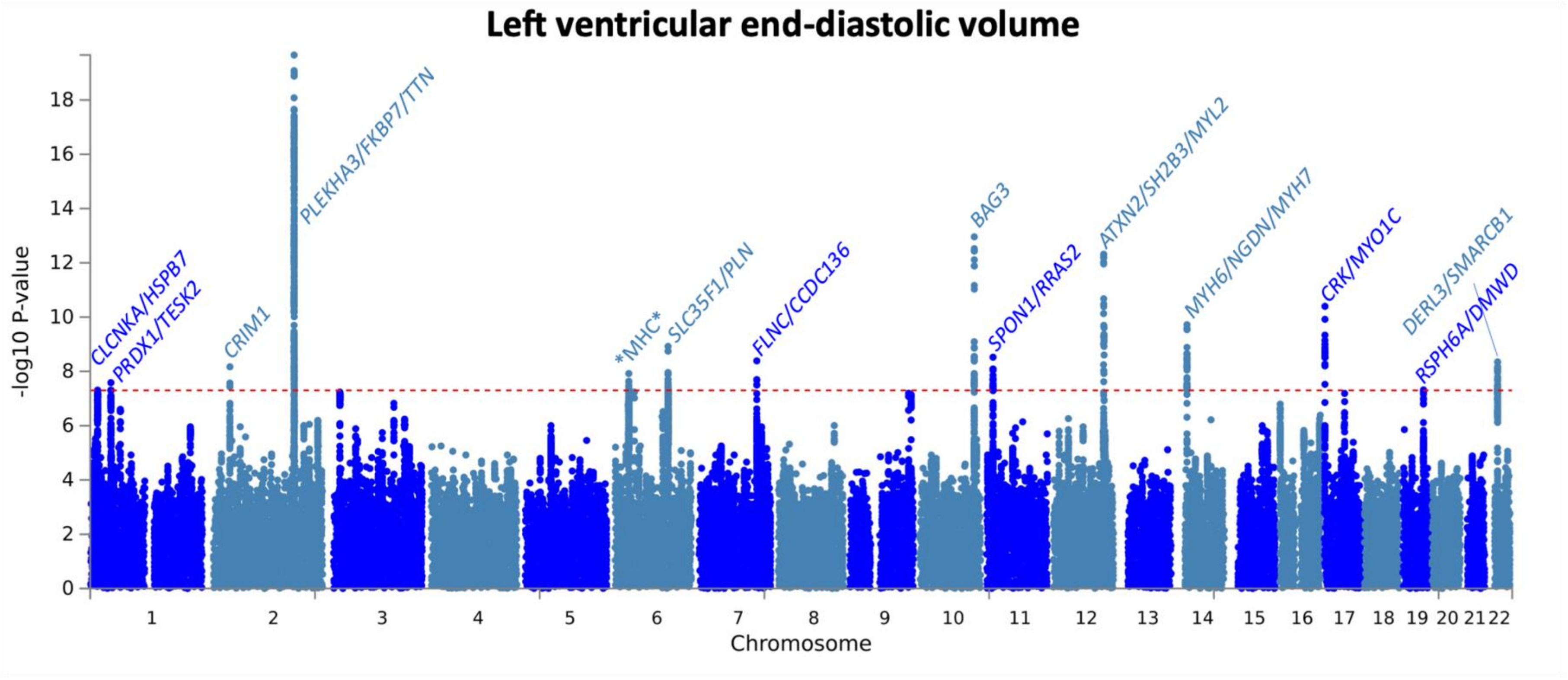


**Supplementary Figure 7:** Manhattan plot for GWAS of left ventricular end-diastolic volume, indexed to the body surface area. GWAS based on an analysis of 36,083 UK Biobank participants without cardiomyopathy and with available cardiac magnetic resonance data. Genomic position presented on the x-axis and -log (P-value) on the y-axis. Manhattan plot generated using FUMA[1] and annotated with locus names based on nearest gene, top gene mapped using OpenTargets[2] variant to gene pipeline (25 February 2022 release), and any mendelian cardiomyopathy gene mapped using FUMA v1.3.8. Locus name and data point colors alternate by chromosome. *MHC* refers to the major histocompatibility locus on chromosome 6. See Supplementary Table 8 for description of the lead variants in the LV trait GWAS and lookup in the HCM GWAS.

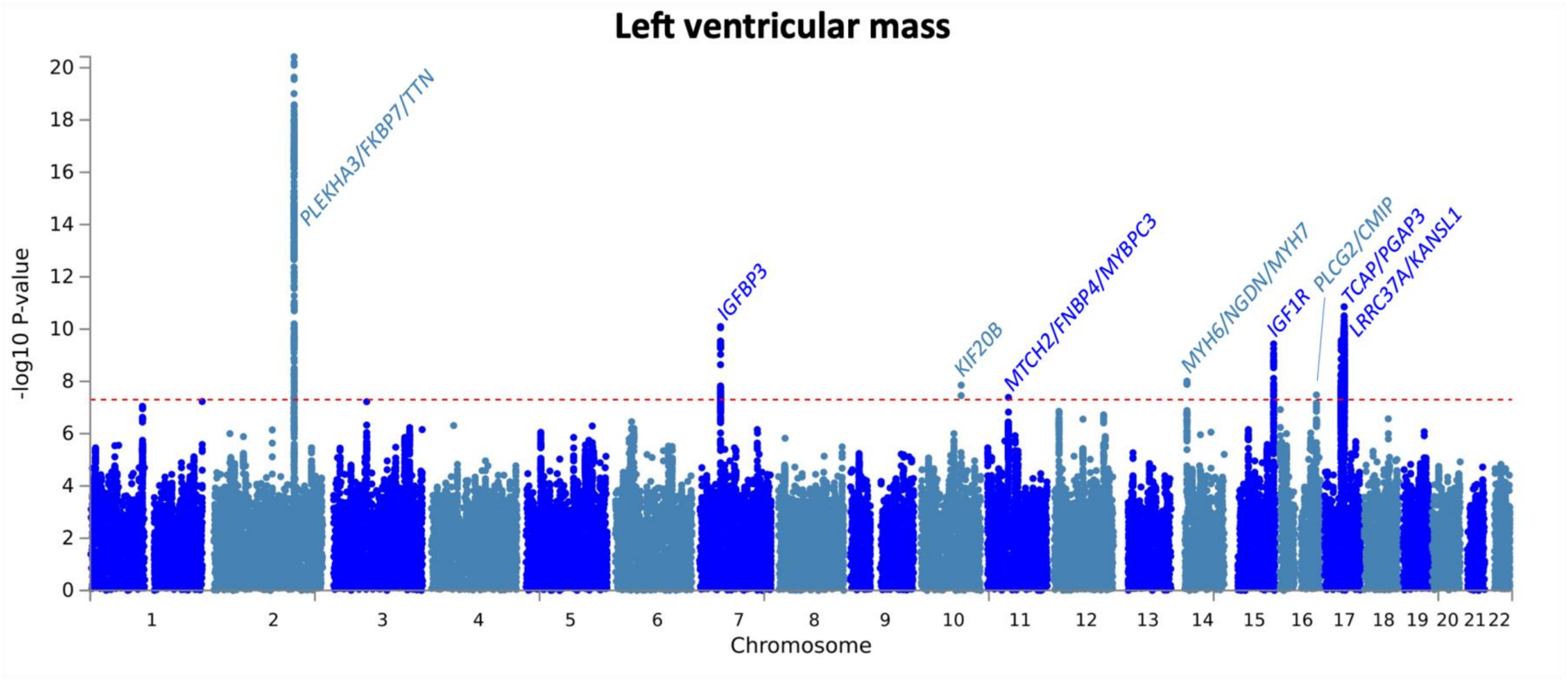


**Supplementary Figure 8:** Manhattan plot for GWAS of left ventricular mass, indexed to the body surface area. GWAS based on an analysis of 36,083 UK Biobank participants without cardiomyopathy and with available cardiac magnetic resonance data. Genomic position presented on the x-axis and -log (P-value) on the y-axis. Manhattan plot generated using FUMA[1] and annotated with locus names based on nearest gene, top gene mapped using OpenTargets[2] variant to gene pipeline (25 February 2022 release), and any mendelian cardiomyopathy gene mapped using FUMA v1.3.8. Locus name and data point colors alternate by chromosome. See Supplementary Table 8 for description of the lead variants in the LV trait GWAS and lookup in the HCM GWAS.

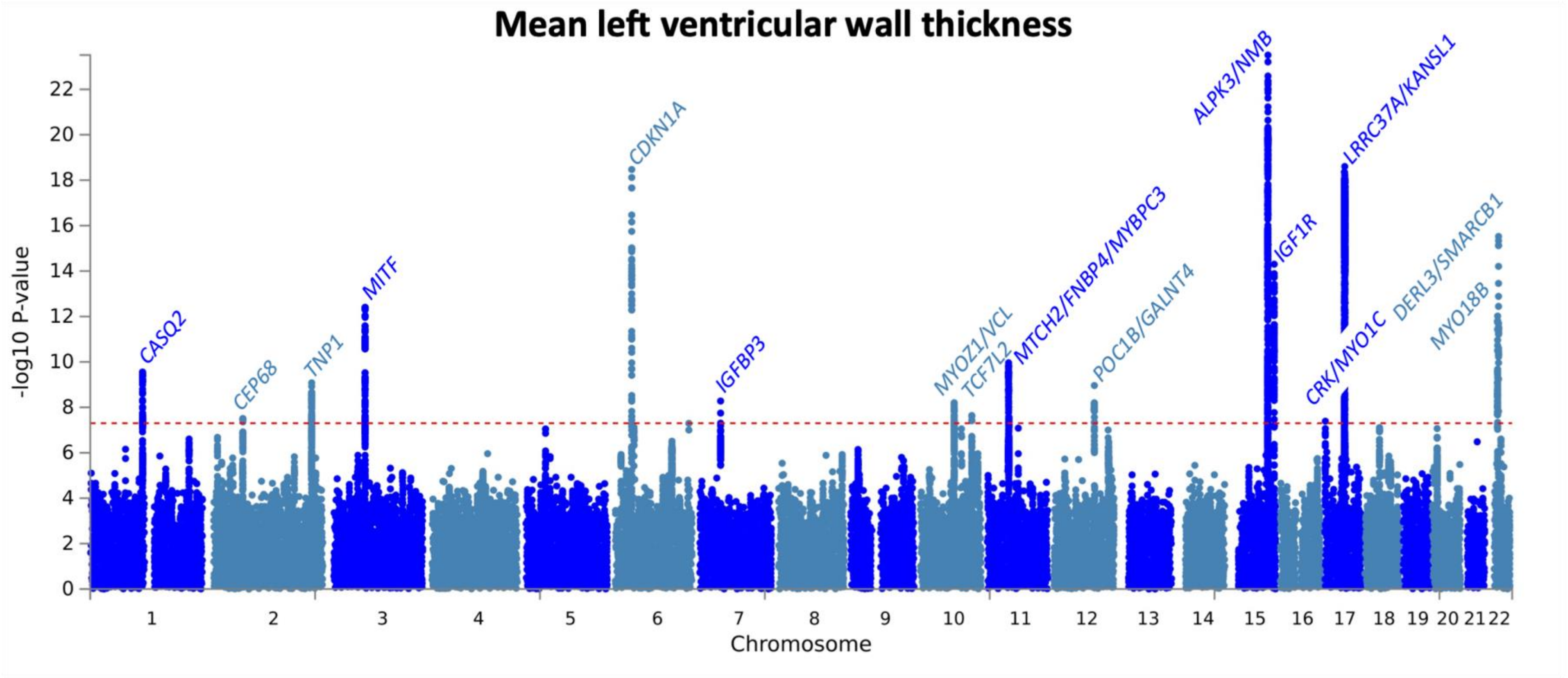


**Supplementary Figure 9:** Manhattan plot for GWAS of mean left ventricular wall thickness. Based on an analysis of 36,056 UK Biobank participants without cardiomyopathy and with available cardiac magnetic resonance data. Genomic position presented on the x-axis and -log (P-value) on the y-axis. Manhattan plot generated using FUMA[1] and annotated with locus names based on nearest gene, top gene mapped using OpenTargets[2] variant to gene pipeline (25 February 2022 release), and any mendelian cardiomyopathy gene mapped using FUMA v1.3.8. Locus name and data point colors alternate by chromosome. See Supplementary Table 8 for description of the lead variants in the LV trait GWAS and lookup in the HCM GWAS.

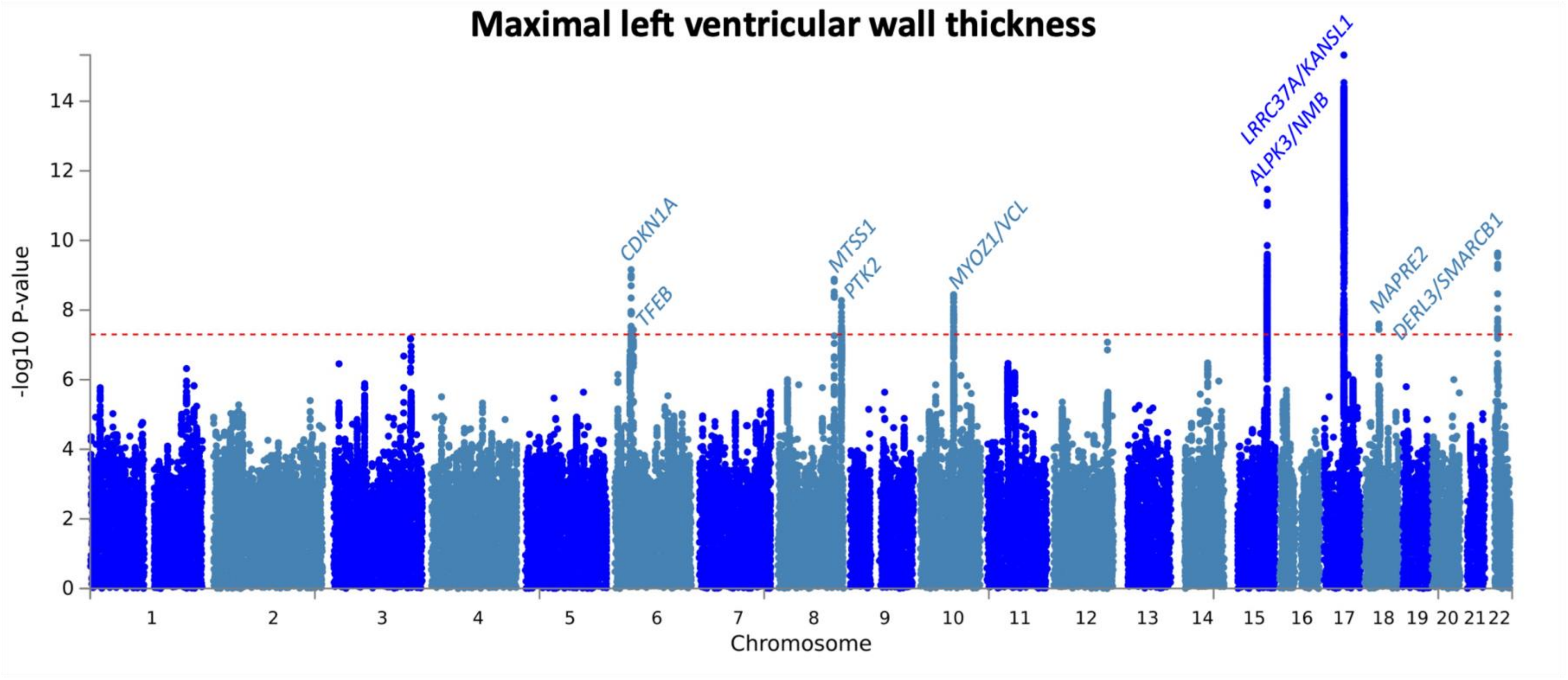


**Supplementary Figure 10:** Manhattan plot for GWAS of maximal left ventricular wall thickness. Based on an analysis of 36,056 UK Biobank participants without cardiomyopathy and with available cardiac magnetic resonance data. Genomic position presented on the x-axis and -log (P-value) on the y-axis. Manhattan plot generated using FUMA[1] and annotated with locus names based on nearest gene, top gene mapped using OpenTargets[2] variant to gene pipeline (25 February 2022 release), and any mendelian cardiomyopathy gene mapped using FUMA v1.3.8. Locus name and data point colors alternate by chromosome. See Supplementary Table 8 for description of the lead variants in the LV trait GWAS and lookup in the HCM GWAS.

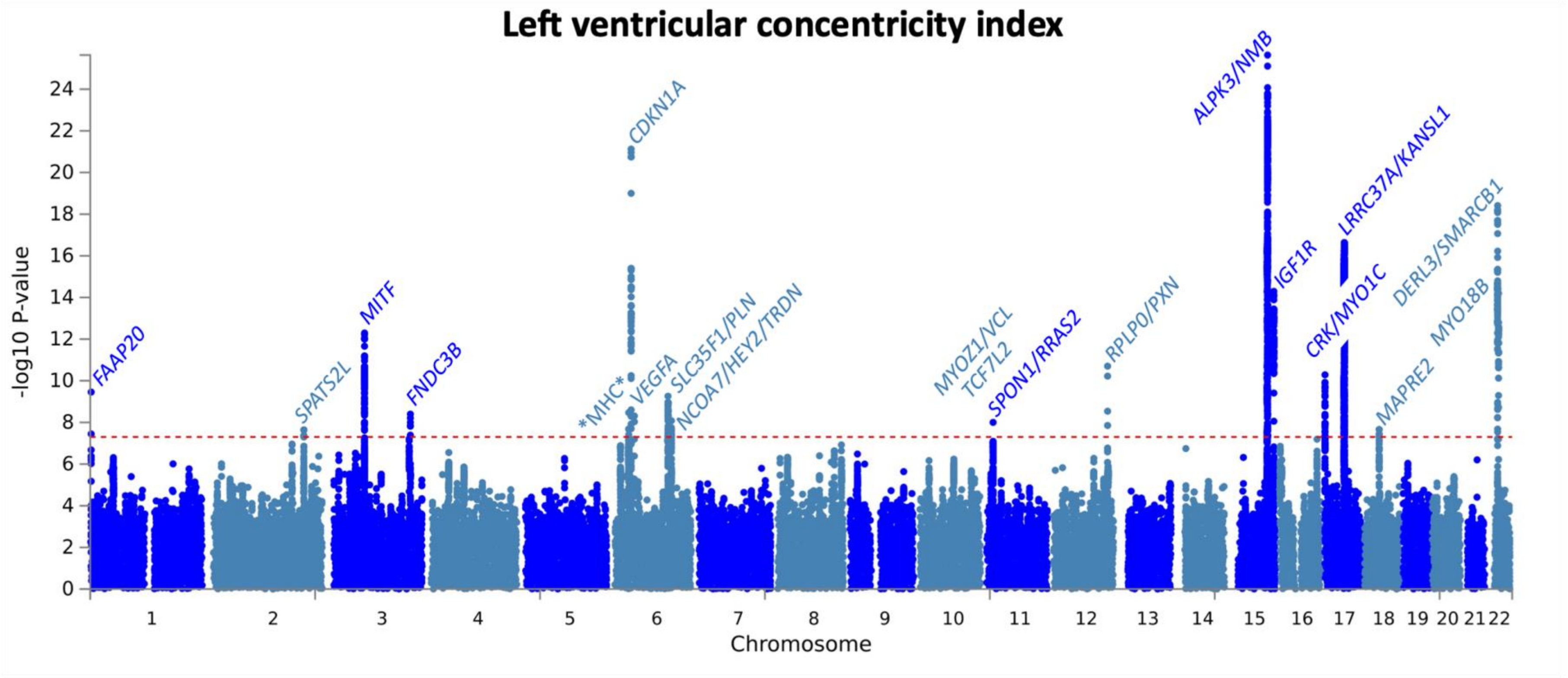


**Supplementary Figure 11:** Manhattan plot for GWAS of left ventricular concentricity index. The left ventricular concentricity index is the ratio of left ventricular mass/left ventricular end-diastolic volume. GWAS based on an analysis of 36,083 UK Biobank participants without cardiomyopathy and with available cardiac magnetic resonance data. Genomic position presented on the x-axis and -log (P-value) on the y-axis. Manhattan plot generated using FUMA[1] and annotated with locus names based on nearest gene, top gene mapped using OpenTargets[2] variant to gene pipeline (25 February 2022 release), and any mendelian cardiomyopathy gene mapped using FUMA v1.3.8. Locus name and data point colors alternate by chromosome. "MHC" refers to the major histocompatibility locus on chromosome 6. See Supplementary Table 8 for description of the lead variants in the LV trait GWAS and lookup in the HCM GWAS.

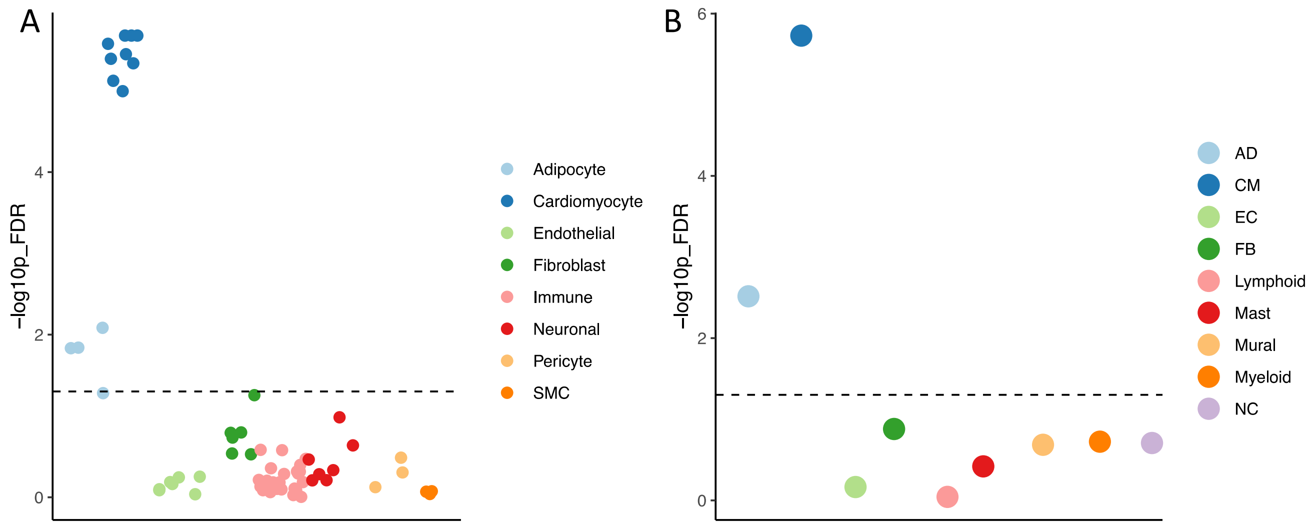


**Supplementary Figure 12**: Enrichment of 68 cell-state (A) and 9 cell-type (B) gene programs from single nuclei data of control heart samples from Reichart D et al[3] using the sc-linker approach and ABC-model enhancer-gene linking strategy. Each point represents a cell state (A) or type (B) with -$\log_{10}$(P) FDR of the heritability enrichment using S-LDSC. Dashed line represents FDR=0.05 for significance.

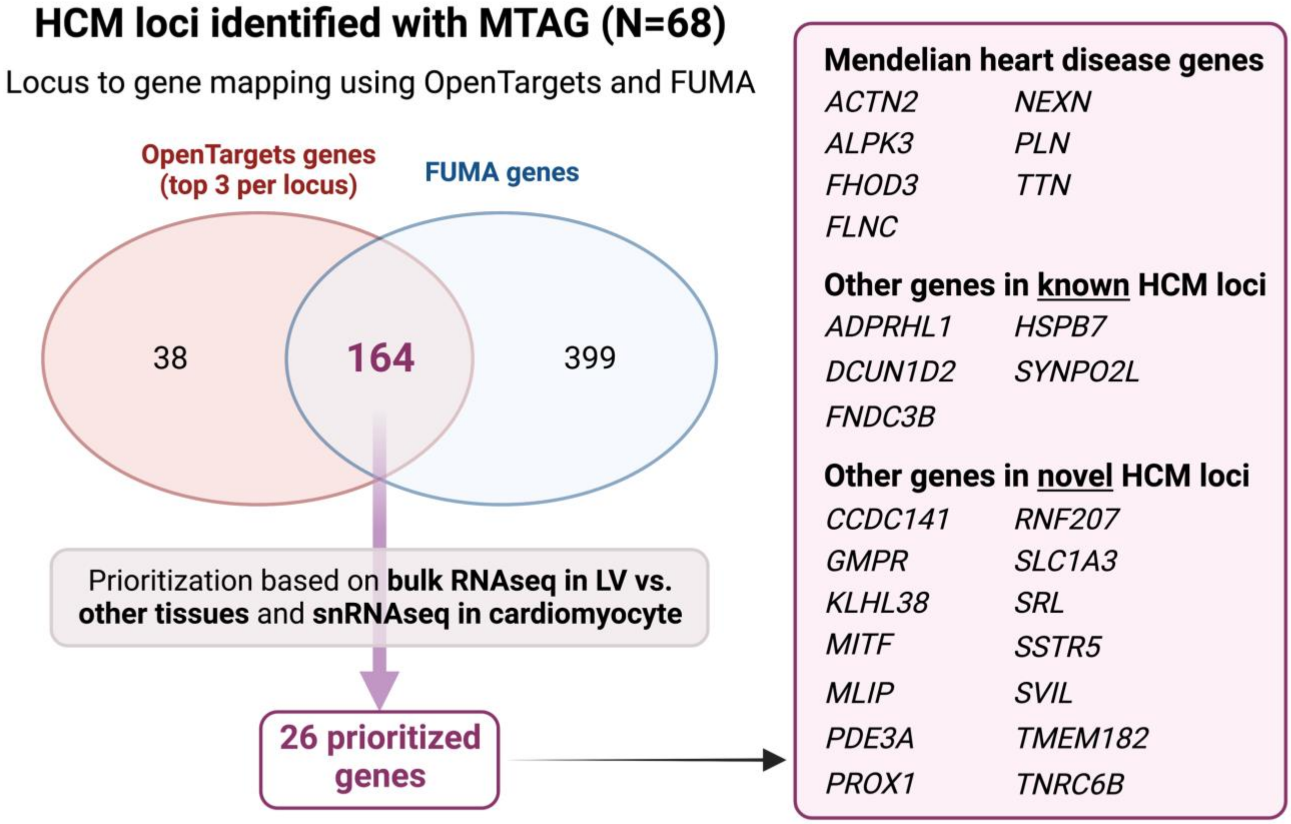


**Supplementary Figure 13**: HCM locus to gene mapping and prioritization based on cardiac expression. Locus to gene mapping was done using the OpenTargets[2] variant to gene (V2G) pipeline (release of 12 October 2022) for all 68 lead variants at the HCM MTAG loci and using FUMA[1] for the HCM MTAG summary statistics (see methods for detailed parameters). From 164 genes mapped using both FUMA and OpenTargets (top 3 genes per locus), 26 were prioritized because of either high specificity of LV expression using the bulk RNAseq data of the Genotype tissue expression (GTEx) project[4] release v8 and/or high expression in cardiomyocytes using snRNA-seq data of Chaffin et al[5] (data accessed using the Broad Institute single cell portal). See methods and Supplementary Tables 13 and 14 for details.

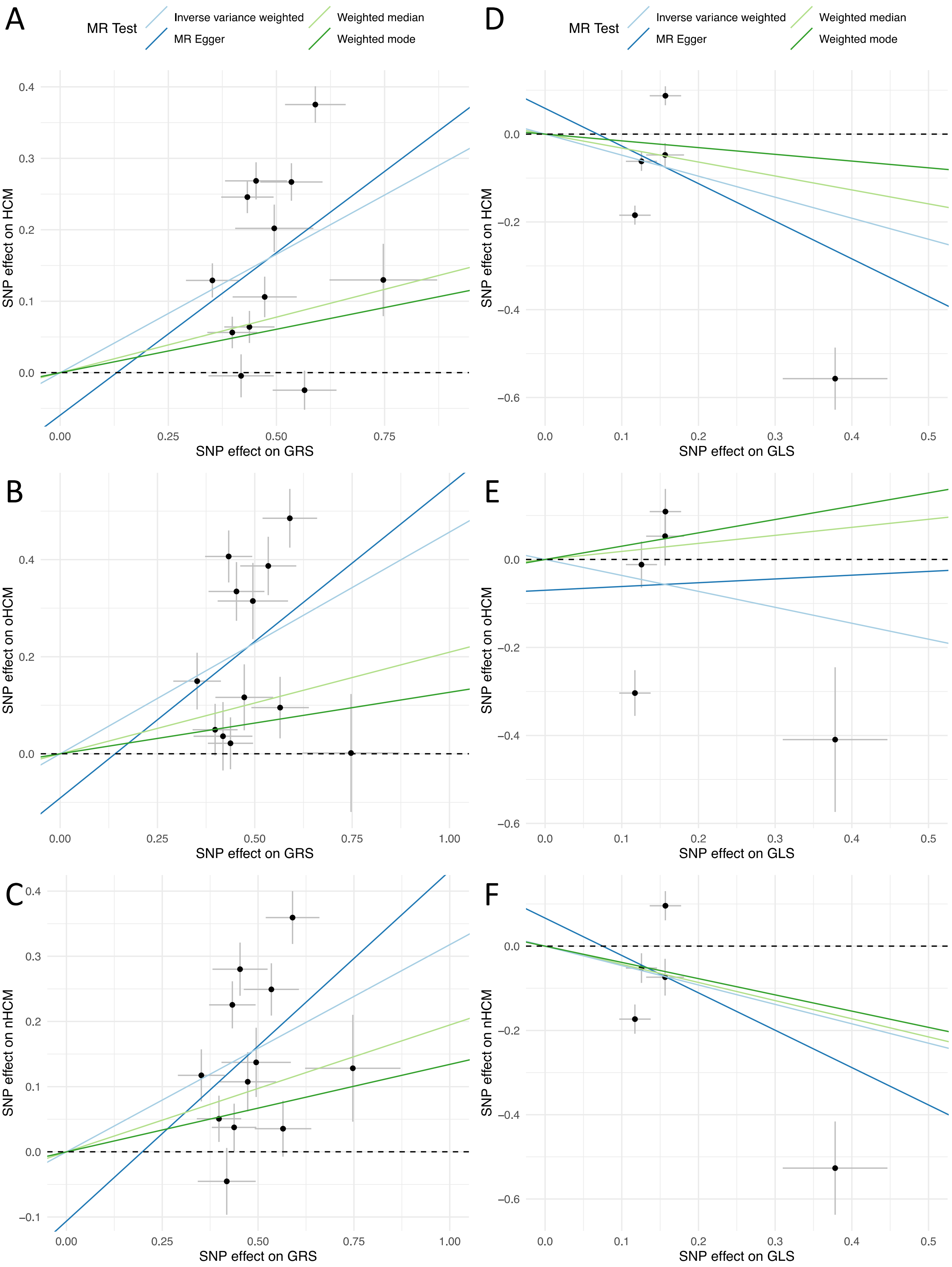
A
MR Test
Inverse variance weighted
MR Egger
Weighted median
Weighted mode
SNP effect on HCM
SNP effect on GRS
D
MR Test
Inverse variance weighted
MR Egger
Weighted median
Weighted mode
SNP effect on HCM
SNP effect on GLS
B
SNP effect on oHCM
SNP effect on GRS
E
SNP effect on oHCM
SNP effect on GLS
C
SNP effect on nHCM
SNP effect on GRS
F
SNP effect on nHCM
SNP effect on GLS

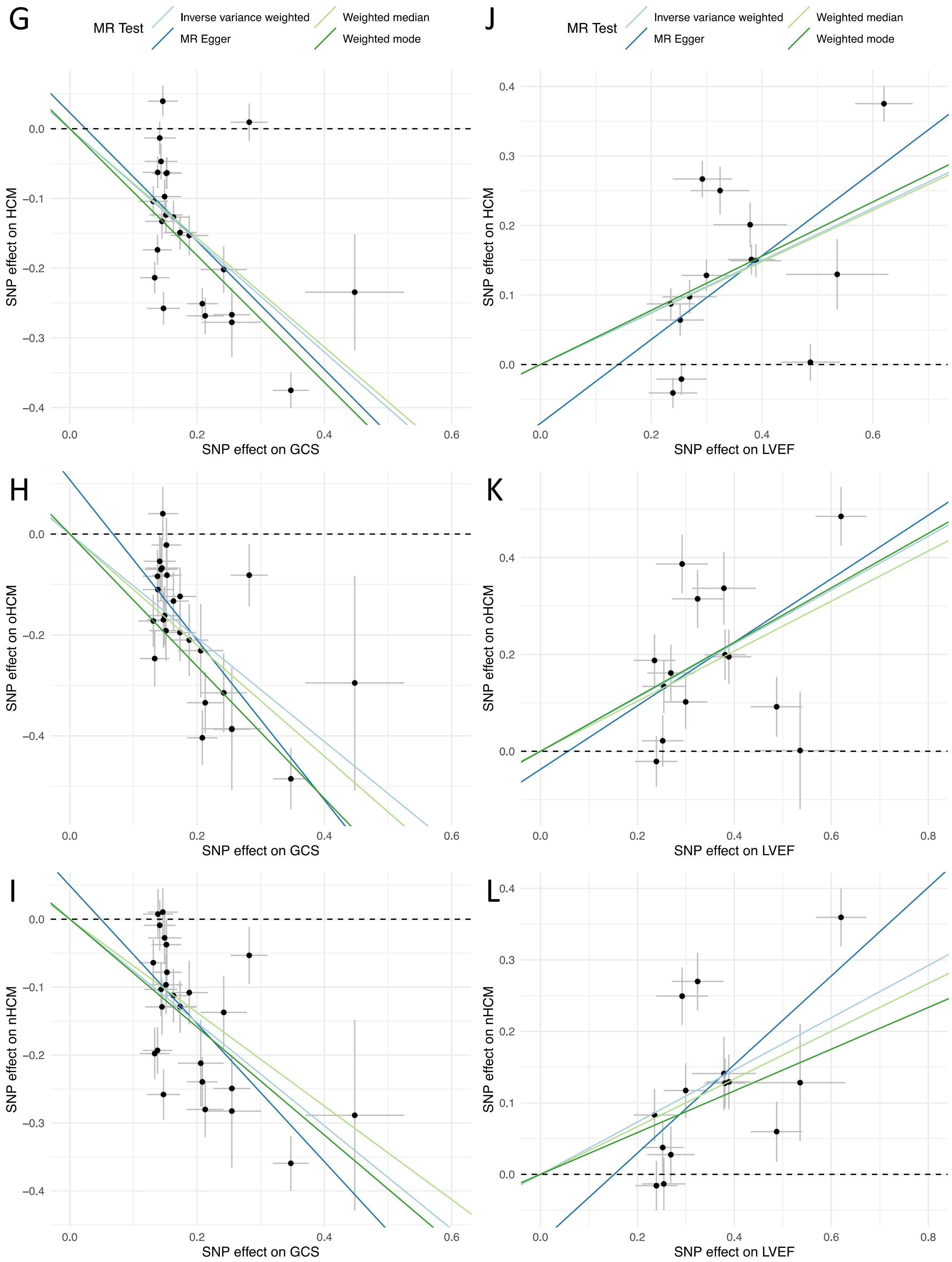
G
MR Test
Inverse variance weighted
MR Egger
Weighted median
Weighted mode
SNP effect on HCM
SNP effect on GCS
J
SNP effect on HCM
SNP effect on LVEF
H
SNP effect on oHCM
SNP effect on GCS
K
SNP effect on oHCM
SNP effect on LVEF
I
SNP effect on nHCM
SNP effect on GCS
L
SNP effect on nHCM
SNP effect on LVEF

↑ **Supplementary Figure 14**: SNP effects of contractility on risk of HCM, oHCM and nHCM. Mendelian randomization (MR) analysis of LV contractility (exposure) on risk of hypertrophic cardiomyopathy (HCM, top panels) and its obstructive (oHCM, middle panels) and non-obstructive (nHCM, bottom panels) forms (outcomes). Genetic instruments for LV contractility were selected from the present GWAS of global strain in the radial (GRS, panels A-C), longitudinal (GLS, panels D-F) and circumferential (GCS, panels G-I) directions, and left ventricular ejection fraction (LVEF, panels J-L), in up to 36,083 participants of the UKB without cardiomyopathy and with available CMR. The outcome HCM GWAS included 5,927 HCM cases vs. 68,359 controls. Of those, 964 cases and 27,163 controls were included in the oHCM GWAS, and 2,491 cases and 27,109 were included in the nHCM GWAS. The effects ($\beta$) of the exposure variable-increasing allele at all independent SNPs reaching $P<5x10^{-8}$ are plotted as datapoints and associated standard errors are represented as lines extending from the datapoints. The x-axis represents the $\beta$ for the exposure variable (contractility: GRS, GLS, GCS, or LVEF) and the y-axis represents the $\beta$ for the outcome variable (HCM, oHCM, or nHCM). Lines represent the causal effects using 4 MR models (legend on top applicable for all panels). Note that a positive $\beta$ for LVEF and GRS represents *increased* contractility, while for GCS and GLS it reflects *decreased* contractility since GCS and GLS are negative measures where increasingly negative values reflect increase in contractility. See Figure 5 and Supplementary Table 16 for full MR results.

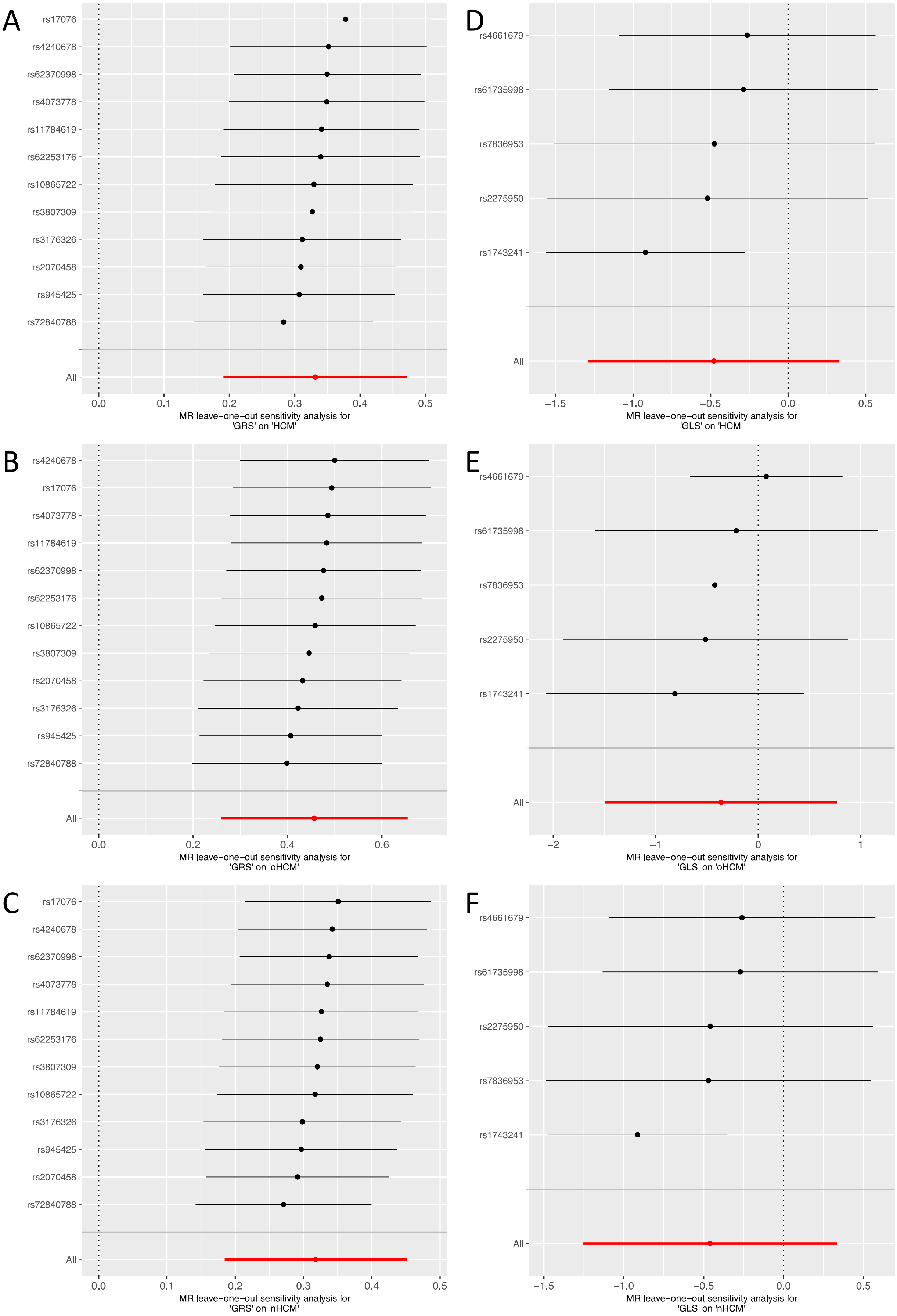
A
rs17076
rs4240678
rs62370998
rs4073778
rs11784619
rs62253176
rs10865722
rs3807309
rs3176326
rs2070458
rs945425
rs72840788
All
0.0
0.1
0.2
0.3
0.4
0.5
MR leave−one−out sensitivity analysis for
'GRS' on 'HCM'
D
rs4661679
rs61735998
rs7836953
rs2275950
rs1743241
All
−1.5
−1.0
−0.5
0.0
0.5
MR leave−one−out sensitivity analysis for
'GLS' on 'HCM'
B
rs4240678
rs17076
rs4073778
rs11784619
rs62370998
rs62253176
rs10865722
rs3807309
rs2070458
rs3176326
rs945425
rs72840788
All
0.0
0.2
0.4
0.6
MR leave−one−out sensitivity analysis for
'GRS' on 'oHCM'
E
rs4661679
rs61735998
rs7836953
rs2275950
rs1743241
All
−2
−1
0
1
MR leave−one−out sensitivity analysis for
'GLS' on 'oHCM'
C
rs17076
rs4240678
rs62370998
rs4073778
rs11784619
rs62253176
rs3807309
rs10865722
rs3176326
rs945425
rs2070458
rs72840788
All
0.0
0.1
0.2
0.3
0.4
0.5
MR leave−one−out sensitivity analysis for
'GRS' on 'nHCM'
F
rs4661679
rs61735998
rs2275950
rs7836953
rs1743241
All
−1.5
−1.0
−0.5
0.0
0.5
MR leave−one−out sensitivity analysis for
'GLS' on 'nHCM'

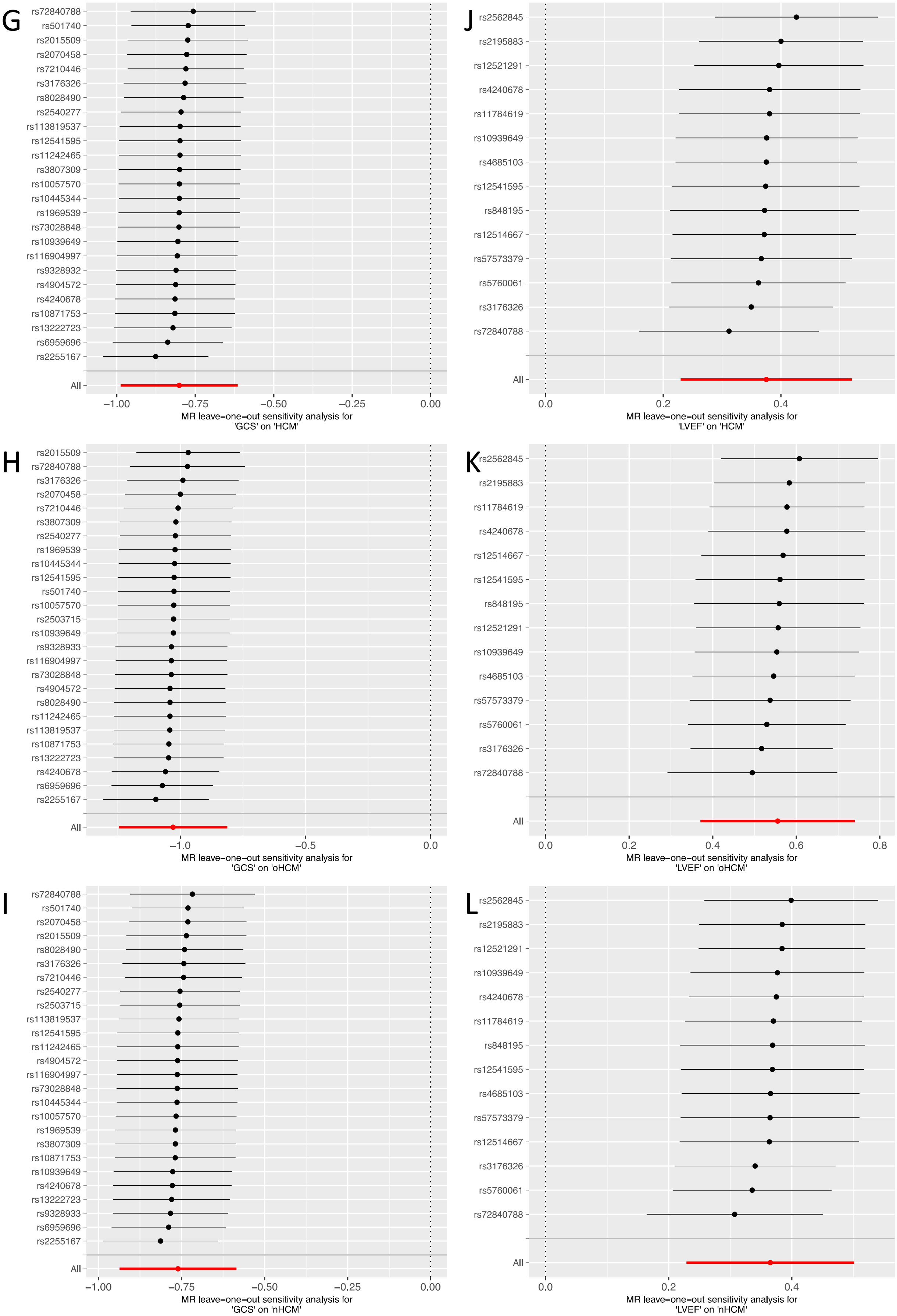
G
rs72840788
rs501740
rs2015509
rs2070458
rs7210446
rs3176326
rs8028490
rs2540277
rs113819537
rs12541595
rs11242465
rs3807309
rs10057570
rs10445344
rs1969539
rs73028848
rs10939649
rs116904997
rs9328932
rs4904572
rs4240678
rs10871753
rs13222723
rs6959696
rs2255167
All
−1.00
−0.75
−0.50
−0.25
0.00
MR leave−one−out sensitivity analysis for
'GCS' on 'HCM'
J
rs2562845
rs2195883
rs12521291
rs4240678
rs11784619
rs10939649
rs4685103
rs12541595
rs848195
rs12514667
rs57573379
rs5760061
rs3176326
rs72840788
All
0.0
0.2
0.4
MR leave−one−out sensitivity analysis for
'LVEF' on 'HCM'
H
rs2015509
rs72840788
rs3176326
rs2070458
rs7210446
rs3807309
rs2540277
rs1969539
rs10445344
rs12541595
rs501740
rs10057570
rs2503715
rs10939649
rs9328933
rs116904997
rs73028848
rs4904572
rs8028490
rs11242465
rs113819537
rs10871753
rs13222723
rs4240678
rs6959696
rs2255167
All
−1.0
−0.5
0.0
MR leave−one−out sensitivity analysis for
'GCS' on 'oHCM'
K
rs2562845
rs2195883
rs11784619
rs4240678
rs12514667
rs12541595
rs848195
rs12521291
rs10939649
rs4685103
rs57573379
rs5760061
rs3176326
rs72840788
All
0.0
0.2
0.4
0.6
0.8
MR leave−one−out sensitivity analysis for
'LVEF' on 'oHCM'
I
rs72840788
rs501740
rs2070458
rs2015509
rs8028490
rs3176326
rs7210446
rs2540277
rs2503715
rs113819537
rs12541595
rs11242465
rs4904572
rs116904997
rs73028848
rs10445344
rs10057570
rs1969539
rs3807309
rs10871753
rs10939649
rs4240678
rs13222723
rs9328933
rs6959696
rs2255167
All
−1.00
−0.75
−0.50
−0.25
0.00
MR leave−one−out sensitivity analysis for
'GCS' on 'nHCM'
L
rs2562845
rs2195883
rs12521291
rs10939649
rs4240678
rs11784619
rs848195
rs12541595
rs4685103
rs57573379
rs12514667
rs3176326
rs5760061
rs72840788
All
0.0
0.2
0.4
MR leave−one−out sensitivity analysis for
'LVEF' on 'nHCM'

↑ **Supplementary Figure 15**: Leave-one-out analyses for contractility Mendelian randomization (MR) of LV contractility on risk of hypertrophic cardiomyopathy (HCM, top panels) and its obstructive (oHCM, middle panels) and non-obstructive (nHCM, bottom panels) forms. Genetic instruments for LV contractility were selected from the present GWAS of global strain in the radial (GRS, panels A-C), longitudinal (GLS, panels D-F) and circumferential (GCS, panels G-I) directions, and left ventricular ejection fraction (LVEF, panels J-L). The IVW MR method is used to estimate the causal effect of contractility on HCM (x-axis) leaving out one SNP at a time (y-axis) to assess whether the association is driven by this single SNP. The IVW effect estimate (center mark) is shown along with the standard error (bars). The overall effect estimate for each contractility measure is shown in the corresponding panel in red. All associations are significant overall and when excluding each SNP at a time, except for GLS.

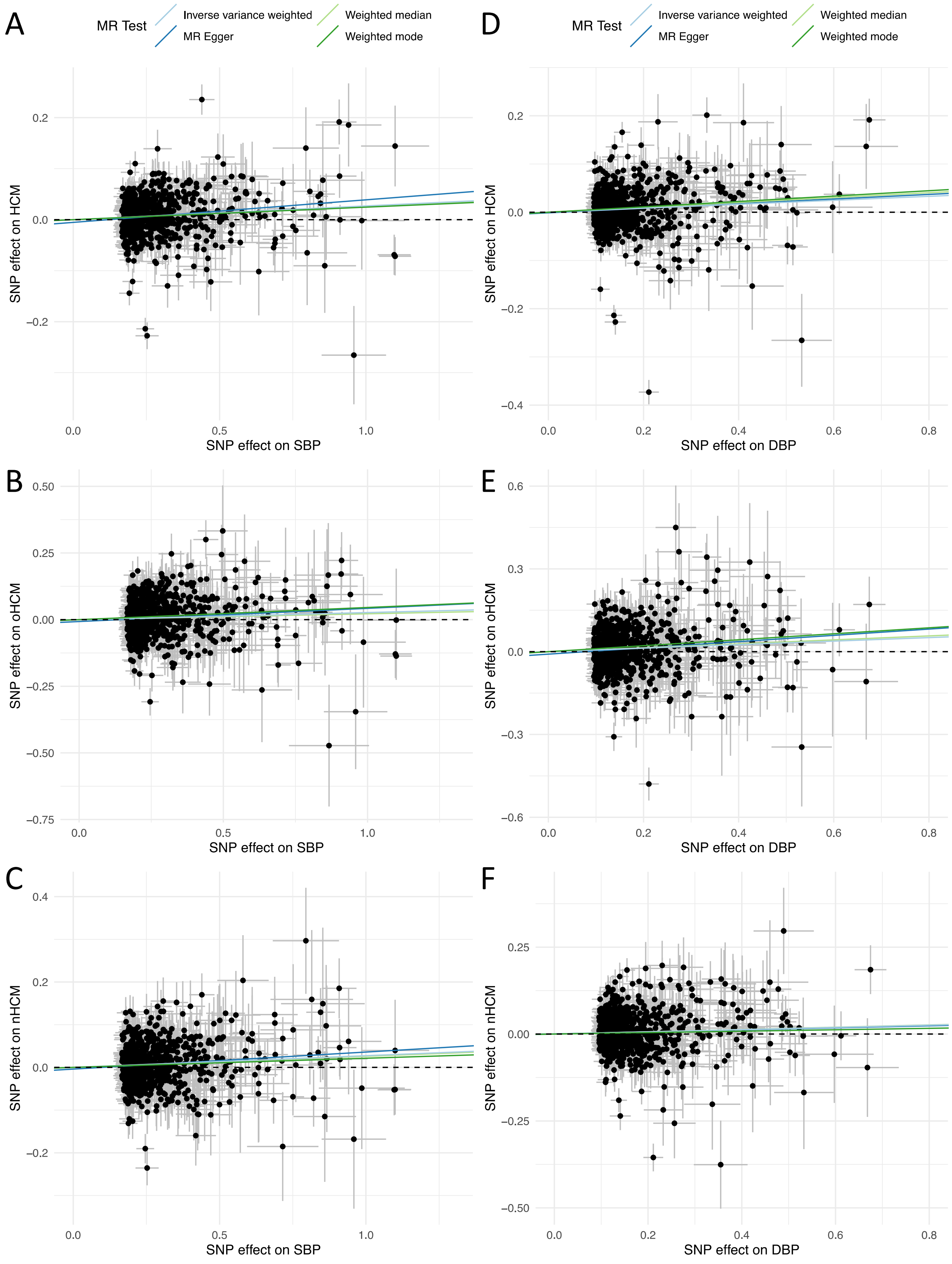
A
MR Test
Inverse variance weighted
MR Egger
Weighted median
Weighted mode
0.2
0.0
−0.2
SNP effect on HCM
0.0
0.5
1.0
SNP effect on SBP
D
MR Test
Inverse variance weighted
MR Egger
Weighted median
Weighted mode
0.2
0.0
−0.2
−0.4
SNP effect on HCM
0.0
0.2
0.4
0.6
0.8
SNP effect on DBP
B
0.50
0.25
0.00
−0.25
−0.50
−0.75
SNP effect on oHCM
0.0
0.5
1.0
SNP effect on SBP
E
0.6
0.3
0.0
−0.3
−0.6
SNP effect on oHCM
0.0
0.2
0.4
0.6
0.8
SNP effect on DBP
C
0.4
0.2
0.0
−0.2
SNP effect on nHCM
0.0
0.5
1.0
SNP effect on SBP
F
0.25
0.00
−0.25
−0.50
SNP effect on nHCM
0.0
0.2
0.4
0.6
0.8
SNP effect on DBP

↑ **Supplementary Figure 16**: SNP effects of blood pressure on risk of HCM, oHCM and nHCM. Mendelian randomization (MR) analysis of systolic (SBP, panels A-C) and diastolic (DBP, panels D-F) blood pressure (exposure) on risk of hypertrophic cardiomyopathy (HCM, top panels A and D) and its obstructive (oHCM, middle panels B and E) and non-obstructive (nHCM, bottom panels C and F) forms (outcomes). Genetic instruments for SBP and DBP were selected from a published GWAS of blood pressure including up to 801,644 individuals.[6] The outcome HCM GWAS included 5,927 HCM cases vs. 68,359 controls. Of those, 964 cases and 27,163 controls were included in the oHCM GWAS, and 2,491 cases and 27,109 were included in the nHCM GWAS. The effects ($\beta$) of the exposure variable-increasing allele at all independent SNPs reaching $P<5x10^{-8}$ are plotted as datapoints and associated standard errors are represented as lines extending from the datapoints. The x-axis represents the $\beta$ for the exposure variable (SBP or DBP) and the y-axis represents the $\beta$ for the outcome variable (HCM, oHCM, or nHCM). Lines represent the causal effects using 4 MR models (legend on top applicable for all panels). See Figure 5 and Supplementary Table 16 for full MR results.

**Supplementary Note**

HCM GWAS study cohorts

The HCM meta-analysis included 7 cohorts, described below and in Supplementary Table 1.

*The Hypertrophic Cardiomyopathy Registry (HCMR)*. Details on the HCMR cohort are described in Harper et al[7] and Neubauer et al[8] Briefly, HCMR recuited 2755 incident HCM cases with evidence of unexplained left ventricular hypertrophy (LV wall thickness, LVWT > 15mm). After QC and removal of cases common to both HCMR and other cohorts described below, 2431 HCM cases were available for analysis. HCM cases underwent gene panel sequencing; variants identified within 8 core sarcomere genes (*MYBPC3*, *MYH7*, *TNNI3*, *TNNT2*, *MYL2*, *MYL3*, *ACTC1* and *TPM1*) were classified using the American College of Medical Genetics and Genomics (ACMG) guidelines.[9] HCM cases were dichotomised into sarcomere-positive (n=850) and sarcomere-negative (N=1,546) groups using a classification framework previously reported in Neubauer et al.[8] An all-comer analysis evaluated 2,431 HCM cases and 40,283, age- and sex-matched, UK Biobank (UKB) participants (application 11223) as controls. Separate sarcomere-positive and sarcomere-negative HCM analyses were performed using randomly allocated non-overlapping 20K UKB controls. Analyses were performed with logistic regression using SNPTEST[10] v2.5.4-beta3 with the Newton-Raphson method adjusting for the first ten ancestry informative principal components (PCs). LD score regression[11] intercept was used to adjust the standard errors for all comer and sarcomere-negative analyses. GC Lambda after correction for both all-comer and sarcomere-negative datasets was 1.05.

*Canadian HCM cohort*. Unrelated cases diagnosed with HCM with included from the Toronto General Hospital HCM clinic, the Montreal Heart Institute (MHI) Cardiovascular Genetics Clinic, the MHI Biobank and the London Health Sciences Centre. All cases had LVWT>15mm or LVWT>13mm in presence of family history of HCM or in presence of a pathogenic or likely pathogenic variant in the 8 core sarcomere genes. Cases were excluded if they had syndromic HCM, metabolic disease, or had >1 sarcomeric pathogenic or likely pathogenic variants (homozygous, compound heterozygous or digenic). Cases underwent targeted sequencing of genes associated with HCM, as per local practice at the time of analysis. Rare variants detected through sequencing in each of the contributing cohorts of this study were assessed centrally for pathogenicity by the Oxford laboratory using the classification framework previously reported in Neubauer et al.[8] Controls without a diagnosis of HCM were included from the MHI Biobank and the London Health Sciences Centre. All cases and controls were genotyped on the Illumina Global Screening Array (GSA). QC was performed as previously described[12], followed by imputation with Minimac4 on the Michigan Imputation server using the Haplotype Reference Consortium r1.1.[13] A total of 1,035 cases and 13,889 controls were then included in the association analysis using SNPTEST v2.5 with the score method adjusting for the first 20 genotypic PCs. In addition to the primary all-comer analysis, a case-control analysis of sarcomere-positive (N=261) and sarcomere-negative (N=582) was performed using random allocation of controls.

*Netherlands HCM cohort*. The Netherlands cohort was previously described.[12] Cases with HCM were included from Amsterdam University Medical Center, Erasmus Medical Center and the University Medical Center Groningen. The control group consisted of a previously genotyped Dutch control population from project Mine (cohort NL4, as described by van Rheenen et al[14]). All cases and controls underwent genome-wide array genotyping on an Illumina Infinium BeadChip. Following QC and imputation, 999 unrelated HCM cases and 2,117 controls were included in the association analysis using SNPTEST v.2.5.2 with the score method correcting for the first 3 PCs. HCM cases were dichotomized into sarcomere-positive (N=436) and sarcomere-negative (N=538) as described for the HCMR cohort and stratified cases control analyses were performed using randomly allocated controls.

*Genomics England 100K Genome Project (GEL).* A GWAS was performed for HCM cases and healthy unrelated controls recruited to the 100K Genome Project (GEL)[15] using whole-genome sequencing data. Cases were defined as individuals recruited to GEL with a primary clinical diagnosis of hypertrophic cardiomyopathy. Controls were selected from healthy relatives recruited to GEL, in whom the proband did not have any cardiac phenotype and matched 5:1 with cases for sex. SNP-level QC excluded variants with a sequencing depth <10, median GQ<15, AB ratio <0.25, missingness >0.05, MAF <0.01, differential missingness between cases and controls $P<10^{-5}$, and Hardy-Weinberg equilibrium test in unrelated controls $P<10^{-5}$. A total of 471 cases and 2,355 controls were included in the association analysis. A mixed-model analysis adjusted for age, sex and the first 10 genotypic PCs was used to account for distant relatedness and population stratification using SAIGE version 0.42.1.[16] All analyses were performed in the Genomics England Research Environment under project ID 291. Since GEL predominantly included sarcomere-negative HCM cases (442/471 cases), no sarcomere-positive analysis was performed.

*Royal Brompton Hospital HCM cohort*. Unrelated British HCM cases from the Royal Brompton & Harefield Hospitals NHS Trust Cardiovascular Research Biobank, and healthy controls from the UK

Digital Heart Project[17] were included. The presence of HCM was excluded using cardiac magnetic resonance (CMR) imaging in all controls. Genotyping was performed in 4 batches at 3 centres (the Sanger Institute, London, Duke-NUS in Singapore and King's college London) using the Illumina Human OmniExpress Beadchip. All downstream QC and data analyses were performed centrally. Data quality control was performed using Plink v1.9 and in-house scripts. We first mapped all SNPs to the positive strand of GRCh37 build. Pre-imputation SNP-level QC excluded SNPs with MAF<0.01, and Hardy-Weinberg equilibrium test $P<10^{-7}$. Sample QC excluded samples with sex mismatch, heterozygosity rate >3 standard deviations (SD) from the mean, and missingness rate >0.05. We aligned study genotypes with those from the HapMap3 cohort, and used PC analysis to identify genotypically Caucasian samples to take forward for imputation, based on cut-offs defined by the mean and SD of each HapMap ethnicity cohort. Batches were merged pre-imputation, excluding SNPs which failed QC checks in any batch. We also checked for differential missingness and excluded any SNPs with differential missingness test $P<1x10^{-7}$. We used the combined UK10K + 1000 Genomes Project dataset as the imputation reference panel. This has a proven record of accuracy for UK study cohorts, across a range of allele frequencies.[18] We pre-phased study genotypes using SHAPEIT[19] (v2.r790) and imputed the genotype data using IMPUTE2[10] (v2.3.2). We finally performed several post-imputation QC steps per-SNP and per-sample, excluding SNPs with an INFO criterion score <0.4, MAF<1%, Hardy-Weinberg equilibrium $P<10^{-7}$. We then removed related individuals from the dataset, using a PI-HAT cut-off of 0.187. After the QC steps, we performed another PC analysis to generate the eigenvectors used as covariates in the regression analysis. Association test was performed by SNPTEST v2.5.4 with the Newton-Raphson method adjusted by sex, age, and top 3 PCs. Stratified analyses for sarcomere-positive (N=118) and sarcomere-negative (N=330) HCM cases using randomly allocated non-overlapping controls were also performed.

*Italian HCM cohort*. Individuals with HCM were diagnosed or referred to the Cardiomyopathy Unit of the Careggi University Hospital (Florence, Italy). HCM diagnosis was defined as unexplained LV hypertrophy with a maximal LV wall thickness >13mm on echocardiography or CMR, complemented by family history and genotype to enable an informed diagnosis. Unrelated healthy Italian controls of European ancestry were obtained from the multicentre international HYPERGENES study.[20] A total of 1,293 control samples were selected, comprising 561 hypertensives (defined as diastolic blood pressure >=90mmHg, systolic blood pressure >=140mmHg, or on antihypertensive treatment before the age of 50) and 732 normotensives. As expected by the inclusion criteria, the mean age was significantly higher in normotensives than in hypertensives. Since aging is associated with an increased prevalence of hypertension, we selected exclusively hyper-normal controls older than 50-year-old allowing for the exclusion of subjects that developed hypertension at a later age. Cases were genotyped at King's College London by Illumina OmniExpress Beadchip and controls were genotyped by Human1M-Duo v3.0 array. All downstream QC and data analyses were performed centrally. Data quality control was performed using Plink v1.9 and in-house scripts. We first mapped all SNPs to the positive strand of GRCh37 build. Pre-imputation SNP-level QC excluded SNPs with MAF<0.01, and Hardy-Weinberg equilibrium test $P<10^{-7}$. Sample QC excluded samples with sex mismatch, heterozygosity rate >3 standard deviations (SD) from the mean, and missingness rate >0.05. We aligned study genotypes with those from the HapMap3 cohort, and used PC analysis to identify genotypically Caucasian samples to take forward for imputation, based on cut-offs defined by the mean and SD of each HapMap ethnicity cohort. Case and control batches were combined after pre-imputation steps. We also checked for differential missingness and excluded any SNPs with differential missingness test $P<1x10^{-7}$. Genome wide imputation was performed using eagle v2.4 phasing, Minimac4 1.5.7 and HRC r1.1 reference panel implemented on the Michigan Imputation Server v1.2.4.[13] We finally performed a number of post-imputation QC steps per-SNP and per-sample, excluding SNPs with a Minimac $R^2<0.5$, MAF<1%, Hardy-Weinberg equilibrium $P<10^{-7}$. We then removed related individuals from the dataset, using a PI-HAT cut-off of 0.187. After the QC steps, we performed another PC analysis to generate the eigenvectors used as covariates in the regression analysis. Association test was performed by SNPTEST version 2.5.4 with the expected method adjusted by age, sex and top 10 PCs. Stratified analyses for sarcomere-positive (N=111) and sarcomere-negative (N=183) HCM cases using randomly allocated non-overlapping controls were also performed. Note that the total number of HCM cases included in the sarcomere status stratified analyses was higher than the number of cases included in the all-comer analysis which was completed at an earlier phase.

*The BioResource for Rare Disease (BRRD)*. The BRRD cohort, a pilot study of the Genomics England 100,000 Genomes Project (GEL), has been described elsewhere.[7,21] In brief, following QC, 239 sarcomere negative HCM cases and 7,203 controls were available for analyses. Logistic regression analysis was performed using SAIGE (v0.29.4.2)[16] with correction for the first three genotypic PCs.

Automated cardiac magnetic resonance (CMR) image analysis

The LV traits GWAS was performed using CMR data from 39,559 UK Biobank participants (36,083 following genotypic and imaging QC and exclusions for co-morbidities). LV trait data were computed using a machine learning analysis of CMR images as described below.

We segmented short and long axis cine images using a fully convolutional network[22], trained on manual annotations of 3,975 subjects, as previously reported.[12] All image segmentations were manually quality controlled by an experienced cardiologist. The segmentation screenshots for short-axis and long-axis end-diastolic and end-systolic frames were visually inspected. Bad segmentations, images with insufficient coverage of the LV or missing anatomical structures were discarded. For motion tracking, subjects with failed image registration or outlier peak global strain values (positive circumferential or longitudinal strain, or negative radial strain) were discarded.

LV end-diastolic (LVEDV) and end-systolic (LVESV) volumes and ejection fraction (LVEF, defined as [LVEDV-LVESV]/LVEDV) were derived from these segmentations. The LV myocardial mass (LVM) was calculated from the myocardial volume using a density of 1.05 g/mL. LV concentricity (LVconc) was defined as LVM/LVEDV. The LV wall thickness (WT) was measured at end-diastole. The myocardium was divided into 16 segments, according to the American Heart Association nomenclature.[23] Maximum and mean wall thickness was derived for each AHA segment. Overall mean wall thickness (meanWT) per subject was calculated from the mean of each of the 16 mean wall thickness measurements. Overall maximum wall thickness (maxWT) was the maximum wall thickness measurement from any segment.

Motion tracking was performed using non-rigid image registration between successive timeframes, using the MIRTK toolkit.[24] Inter-frame displacement fields were composed to obtain the displacement with respect to a reference frame (the end-diastolic frame, or frame 0). To avoid drift effect due to accumulation of registration errors[25], motion tracking is performed twice – along the forward direction (tracking starting from frame 0 to frames 1, 2, 3,…) and backward direction (tracking from frame 0 to frames $T$-1, $T$-2, $T$-3, …) where $T$ denotes the total number of frames per cardiac cycle). The average displacement field is calculated by weighted averaging of the forward and backward displacement field such that, for a frame at the start of the cardiac cycle, the forward displacement field will have a higher weight, whereas for a frame at towards the end of the cardiac cycle, the backward displacement field will have a higher weight. Four image slices were used for this: in the short axis, a basal slice at 75% LV location, a mid-cavity location and an apical slice at 25% LV location, and in the long axis, a 4-chamber view as previously described.[26,27] The myocardial contours on the three slices were divided into the 16 segments. Based on the displacement field from motion tracking, myocardial contours at the end-diastolic frame were warped onto each time frame of the cardiac cycle. Circumferential, radial and longitudinal strains were calculated for each time frame based on the change of length for each line segment[28], using the equation $E = \frac{\Delta L}{L}$, where $E$ is the strain, $\Delta L$ is the change in length of the line segment, and $L$ is the starting (end-diastolic) length of the line segment. Peak strain for each AHA segment, and *global* peak strain were calculated in radial ($strain^{rad}$), longitudinal ($strain^{long}$) and circumferential ($strain^{circ}$) directions.[28] Note that in contrast to $strain^{rad}$, both $strain^{long}$ and $strain^{circ}$ are negative values where more negative values reflect higher contractility. As such, we will sometimes present results for $-strain^{long}$ and $-strain^{circ}$ to facilitate interpretation of directionality.

LV traits GWAS: genotyping and imputation

Details of the genotyping and QC strategy employed by UKB were previously published.[29] Genotypes were called from 2 purpose-built arrays: the UK Biobank Axiom Array (825,927 markers) and UK BiLEVE Axiom Array (807,411 markers). These directly called genotypes were imputed to more than 90 million variants using the haplotype reference consortium (HRC) and the UK10K reference panels. See UKB website for details of imputation and genotype quality control performed by a collaborative group headed by the Wellcome Trust Centre for Human Genetics. We excluded samples with outlying heterozygosity or missingness rates, as defined by UKB and those with mismatches between the genotypic and recorded sex, as well as those with aneuploidy or excess kinship, as defined by UKB. We excluded SNPs failing UKB protocols (filtered per batch by Hardy-Weinberg equilibrium and missingness)[29], those with imputation INFO score <0.3 or MAF <0.01, or with missingness >0.1.
